# Vocal and musical emotion perception, voice cue discrimination, and quality of life in cochlear implant users with and without acoustic hearing

**DOI:** 10.1101/2024.04.28.24305905

**Authors:** Eleanor E. Harding, Etienne Gaudrain, Barbara Tillmann, Bert Maat, Robert L. Harris, Rolien H. Free, Deniz Başkent

**Author notes:** **Corresponding author:** Eleanor E. Harding Department of Otorhinolaryngology, University Medical Center Groningen Hanzeplein 1 9713 GZ, Groningen, The Netherlands.

## Abstract

This study aims to provide a comprehensive picture of auditory emotion perception in cochlear implant (CI) users by (1) investigating emotion categorization in both vocal (pseudo-speech) and musical domains and (2) how individual differences in residual acoustic hearing, sensitivity to voice cues (voice pitch, vocal tract length), and quality of life (QoL) might be associated with vocal emotion perception and, going a step further, also with musical emotion perception. In 28 adult CI users, with or without self-reported acoustic hearing, we showed that sensitivity (d’) scores for emotion categorization varied largely across the participants, in line with previous research. However, within participants, the d’ scores for vocal and musical emotion categorization were significantly correlated, indicating similar processing of auditory emotional cues across the pseudo-speech and music domains and robustness of the tests. Only for musical emotion perception, emotion d’ scores were higher in implant users with residual acoustic hearing compared to no acoustic hearing. The voice pitch perception did not significantly correlate with emotion categorization in either domain, while the vocal tract length significantly correlated in both domains. For QoL, only the subdomain of Speech production ability, but not the overall QoL scores, correlated with vocal emotion categorization, partially supporting previous findings. Taken together, results indicate that auditory emotion perception is challenging for some CI users, possibly a consequence of how available the emotion-related cues are via electric hearing. Improving these cues, either via rehabilitation or training, may also help auditory emotion perception in CI users.

## Introduction

Cochlear implants (CIs) are prosthetic devices that provide hearing to individuals with severe or profound deafness by directly stimulating the auditory nerve using electric current. However, due to the properties of electrode-nerve interface, the transmitted information is reduced in spectrotemporal details (for an overview see Başkent et al., 2016). In particular, the perception of vocal and musical emotion is consistently reported to be challenging for CI users (Ambert-Dahan et al., 2015; Chatterjee et al., 2015, 2023; Lin et al., 2022; Luo et al., 2007; Paquette et al., 2018; Volkova et al., 2013). The ability to perceive vocal emotions in CI users has been shown to be linked to voice cue discrimination thresholds for emotion-related acoustic features such as the voice cues of voice pitch (related to voice fundamental frequency, F0) (Barrett et al., 2020; Deroche et al., 2016; Lin et al., 2022) and vocal tract length (VTL; related to formants, Chuenwattanapranithi et al., 2008; Nussbaum et al., 2022; von Eiff et al., 2022) as well as to listeners’ ratings of quality of life (QoL) (Luo et al., 2018). These links have not yet been established for musical emotion perception. Understanding whether the benefit from the CI is comparable for vocal and musical emotion could inform research on optimal processor settings for speech and music as well as help improve audiological assessment and develop new training strategies. For example, training for the perception of generic auditory perceptual features might benefit emotion perception in speech or music (see Fuller et al., 2018). The current study therefore investigated within-participant vocal and musical emotion perception in CI users, and whether vocal or musical emotion perception was associated with voice cue discrimination or QoL. Moreover, due to earlier referrals to CI teams when hearing aids become not sufficiently functional and the relaxing CI candidacy indications, in recent years, more and more CI users have usable residual (low-frequency) acoustic hearing available to them on the contralateral (non-implanted) ear, although the degree of it varies (e.g., Nyirjesy et al., 2023). In this study, we also explored the extent to which self-reported acoustic hearing, when added to CI electric hearing, may affect vocal and music emotion perception. Thus the current study aims to provide a comprehensive picture of auditory emotion perception by CI users, as assessed in two domains, vocal and musical, and in relation to potentially relevant acoustical factors, such as acoustic hearing and voice cue perception, as well as quality of life.

### Vocal and musical emotion perception

Perception of emotions not only relies on the analysis of the relevant acoustic cues, but also the correct interpretation of the (important or life-saving) emotion so that an appropriate response can be given (e.g., fight, flight, or freeze). Despite the complexity of the neural processing of emotions (e.g., Phillips et al., 2003), historically, emotions and their perception have been considered as discrete categories (e.g., Hess, 2017; Hoemann et al., 2020). As a result, paradigms investigating emotion perception use categories to which a stimuli set belongs, and participants are required to assign an emotion category to each of the facial, vocal or musical stimuli (e.g., Banse & Scherer, 1996; Gelder et al., 2010; Pralus et al., 2020; Vieillard et al., 2008). Dimensional models of emotions suggest that emotion in general can be defined along arousal and valence dimensions and their combinations (Russell, 1980; Schlosberg, 1952). Arousal refers to the intensity of affective experience, whether the emotion is stimulating or relaxing, while valence refers to hedonic experience, whether the emotion is positive or negative (Feldman, 1995). Emotion categories in turn can be located in the two-dimensional arousal-valence emotion framework used. Figure 1 shows how participants previously have scaled the dimensions among a pool of 28 emotions (Russell, 1980).

**Figure 1.**
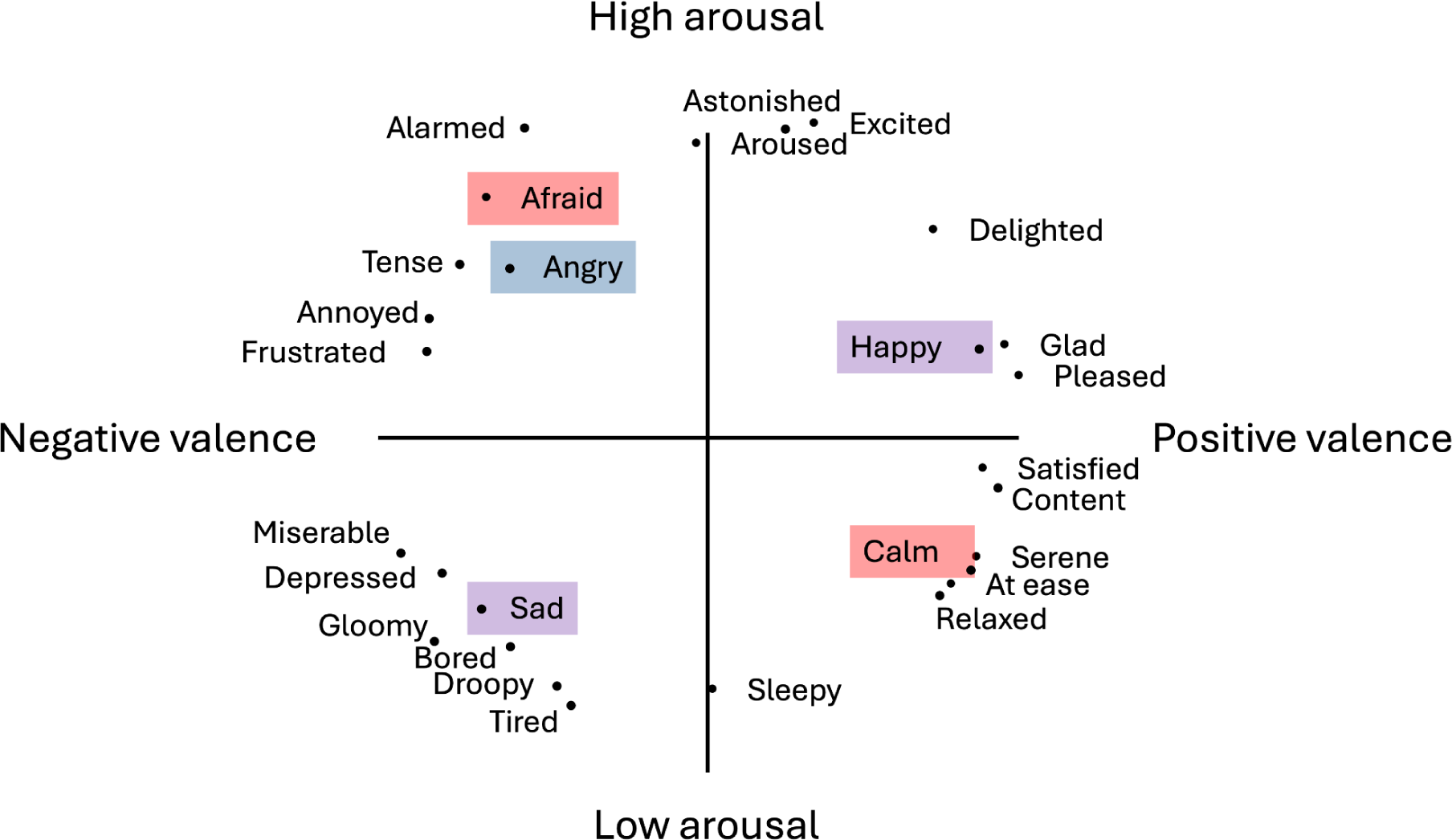
Multidimensional scaling of words describing emotions along arousal and valence dimensions, adapted from Russel (1980). Emotion categories used in this study are highlighted for Experiment 1 (blue), Experiment 2 (red), or both experiments (purple).

#### Acoustic features of vocal emotion

For vocal emotions, arousal has been reported to be expressed by voice intensity, spectral energy content, voice F0, and pitch variability range. High arousal emotions, such as fear, anger, and happiness, are characterized by high intensity and energy, high mean F0, and large F0 variability. In comparison, low-arousal emotions, such as sadness and depression, are characterized by lower energy and lower and less variable F0 (e.g., Banse & Scherer, 1996; Belin et al., 2008). Regarding valence, mean F0 and F0 contour —whether F0 rises or falls across the utterance— could contribute to distinguishing valence in high-arousal emotions (e.g., happy and fearful), though the direction of mean- and F0 contour changes differs between studies and individual speakers (e.g., Bachorowski & Owren, 1995; Belin et al., 2008).

#### Perception of vocal emotions with cochlear implants

When tested for the perception of vocal emotions, CI users typically categorize emotions above chance-level, but display categorization accuracy significantly lower than that of normal-hearing (NH) control participants (Chatterjee et al., 2015, 2023; reviewed in Jiam et al., 2017; Lin et al., 2022; Luo et al., 2007; Paquette et al., 2018; Volkova et al., 2013). Emotion categories are sometimes systematically confused with each other, especially when the categories are located on the same end or half-plane of the arousal dimension, for example happy with scared (e.g., Chatterjee et al., 2015; Lin et al., 2022). Confusions are also reported with a baseline-neutral condition, for example angry and neutral or sad and neutral (e.g., Chatterjee et al., 2015; Luo et al., 2007), though confusion categories can change with stimuli-speaker gender.

#### Acoustic features of musical emotions

For musical emotions, acoustic features have been linked to arousal and valence perception (e.g., Schubert, 2004), and a direct parallel with emotion perception in voice/speech has been proposed (Balkwill & Thompson, 1999). Arousal tends to be expressed by slow-varying temporal cues available in the acoustic envelope of the music, such as perceived loudness changes or how fast or slow the tempo of the beat is (e.g., faster tempi are more stimulating than slower tempi) (e.g., Ilie & Thompson, 2006; Schubert, 2004). Valence, in contrast, is expressed by multifaceted cues conveying the harmonic complexity of the music as well as the pitch intervals governing the mode (Gomez & Danuser, 2007). Within mainstream Western musical culture^1^, major mode and consonance are considered to have a more positive valence (“happy”), whereas minor mode and dissonance indicate more negative valence (“sad or nostalgic”) (e.g., Bigand et al., 1996; Nieminen et al., 2012). Modes are defined according to the intervals between tones of the scale or the melodic contour at structurally important positions, for example “major” and “minor” are different in the harmonic ratio of one interval (the third) changing between 4:5 (major) and 5:6 (minor). Music is considered increasingly dissonant as the ratio between simultaneous or consecutive pitches becomes more complex (e.g., Berg & Stork, 2004), for example, a single note sounding in perfect unison creates a ratio of 1:1, whereas black and white neighboring keys on the piano played together can create a 15:16 ratio (Müller, 2021).

#### Perception of musical emotions with cochlear implants

The perception of musical emotion remains challenging for CI users due to the limited transmission of spectrotemporal information (Caldwell et al., 2017; Giannantonio et al., 2015; Lassaletta et al., 2008; Limb, 2006). Behavioral studies have indicated that perception of cues related to pitch, timbre, harmony, and spectral resolution are limited in CI hearing (Friesen et al., 2001; McDermott, 2004; Shannon, 1983; Zimmer et al., 2019), while the perception of slow-varying temporal information and rhythm seems to be relatively preserved in comparison, though not always unimpaired (Hidalgo et al., 2021). Studies investigating emotion categorization in music have reported that CI users rely more strongly on temporal cues than on pitch cues. A series of studies systematically varied the tempo and mode of melodies with chordal accompaniment and asked CI users to categorize stimuli as happy or sad (Caldwell et al., 2015; Giannantonio et al., 2015; Hopyan et al., 2016). Across the studies, emotion categorization made only with CI hearing (with no additional acoustic hearing) or its simulations (NH children or adults tested with vocoded stimuli as an acoustic simulation of CIs) were based on tempo- and not modal cues.

When not assessing emotion categorization directly, but rather how CI users rate arousal or valence dimensions, CI users’ valence perception more closely resembled NH valence perception than did arousal perception (Ambert-Dahan et al., 2015; Paquette et al., 2018). In one study presenting short bursts of notes played on the violin or clarinet to express the emotion categories happy, fear, sad, and neutral (Paquette et al., 2018), NH listeners appeared^2^ to rate the presence of arousal and valence for items in each emotion category according to their position in the arousal-valence quadrants (see Figure 1). CI users diverged from NH participants in arousal ratings (happy and sad were judged as exhibiting higher arousal than fear and neutral) whereas their valence ratings more closely resembled those of NH participants (happy had more positive valence than fear, but not sad, and fear was marginally more negative than sad). An explanation for improved valence perception compared to arousal perception that was observed in this study could be that the rating of single-instrument note bursts might diverge from polyphonic excerpts used in most other studies or that cues in longer, more complex stimuli include tempo and dynamic changes, both used to encode emotional arousal (Schubert, 2004).

In the other study, musical emotion categorization of classical music excerpts was tested in NH participants and CI users using the emotion categories happiness, fear, sadness and peacefulness (Ambert-Dahan et al., 2015). Additionally, all participants rated the degree of arousal and valence for each emotion category. The arousal ratings were significantly different between NH and CI groups, whereas there was no significant difference in valence ratings. While it remains unclear why arousal perception was more different from NH categorization, this study raises an important point about CI user participants: 11 out of 13 CI users had some usable low-frequency acoustic hearing (enhanced with a contralateral hearing aid) in addition to the electric hearing via their CI, thus some acoustic hearing and acoustic cues were still available. The presence of acoustic hearing in addition to the electric hearing can improve music perception (Fata et al., 2009; Gfeller et al., 2006). This could be due to the rich pitch and harmonic cues available via the low-frequency acoustic hearing.

#### Individual differences in acoustic hearing, voice cue perception, and quality of life

##### Acoustic hearing

In recent years, more and more CI users have usable acoustic hearing available to them, although the degree of it varies (e.g., Nyirjesy et al., 2023). This is due to several interacting factors (and see Snel-Bongers et al., 2018): CI candidacy indications have been evolving; the treatment decision of which ear to implant has shifted to the ear with the worse (as opposed to the better) hearing, leaving more acoustic hearing in the contralateral ear; CI surgeons are trained to use soft surgery skills in all patients with the goal of causing as little trauma in the cochlea as possible and preserving as much residual hearing as possible; CI manufacturing companies are continuing to develop leaner and increasingly atraumatic electrodes. A CI population shift towards more residual acoustic hearing may have consequences for the perception of speech and music. Regarding voice perception, vocoder simulations with additional acoustic hearing limited to 150 or 300 Hz were shown to improve voice pitch perception, indicating that even very limited bandwidth may still provide significant benefits (e.g., Başkent et al., 2016). In contrast, some other studies with CI users did not observe an improvement in within- and and between-gender voice discrimination, where not only voice pitch but other voice characteristics would also differ (Dorman et al., 2008). Music appreciation is also influenced by whether the CI users have usable low-frequency acoustic hearing in the implanted (Gfeller et al., 2006) or non-implanted ear (Fata et al., 2009). Studies have shown that bimodal CI users were able to use pitch-related musical cues to categorize musical emotion more strongly than CI users with no acoustic hearing (D’Onofrio et al., 2020; Giannantonio et al., 2015).

##### Voice cue perception

CI users may be hindered in their perception of emotion categories and, in particular, of the valence dimension by their limited perception of acoustic details. In particular, perception of voice cues of F0 and VTL seems challenging (e.g., Deroche et al., 2014; Gaudrain & Başkent, 2018). F0 is directly related to the glottal pulse rate of the speaker. VTL is strongly associated with speaker’s height (Fitch & Giedd, 1999), with constraints on speech formant frequencies, and is connected to the notion of timbre (Patterson et al., 2010). F0 sensitivity was previously observed to correlate with emotion categorization in CI children (Barrett et al., 2020; Deroche et al., 2016; Lin et al., 2022). VTL (Chuenwattanapranithi et al., 2008), formants (Banse & Scherer, 1996), or more generally, spectral envelopes (Nussbaum et al., 2022; von Eiff et al., 2022) have been shown to contribute to vocal emotion perception in both NH and CI listeners. In CI listeners, VTL discrimination could serve as a proxy measure for spectral resolution (for example, as it seems to relate to other measures, such as perception of speech in speech maskers; El Boghdady et al., 2019). Hence, VTL sensitivity could reflect the ability to perceive the acoustic details encoding valence.

In music, valence perception seems to mostly rely on the perception of major and minor modes, which cue positive or negative valence, respectively. The major/minor distinction has been studied alongside the broader concept of consonance and dissonance, which is evaluated through measures of “pleasantness.” CI users have shown some deficit compared to NH listeners in this area, either with melodies (Caldwell et al., 2016) or with chords (Knobloch et al., 2018). This deficit is likely to find at least some of its origin in the same sensory limitations as the ones that are captured by F0 and VTL discrimination thresholds. If F0 and VTL discrimination thresholds are an overall indication of sound transmission fidelity in CIs, the degree to which these mechanisms are transmitted by the implant may affect both arousal and valence perception.

##### Quality of life

Among CI users, the degree of self-reported QoL was previously reported to be positively associated with vocal emotion perception scores (Luo et al., 2018; Schorr et al., 2009; Von Eiff et al., 2022). One questionnaire specifically developed for CI users is the Nijmegen Cochlear Implant Questionnaire (NCIQ; Hinderink et al., 2000), which assesses hearing-related QoL in the general domains of Physical-, Psychological- and Social functioning. NCIQ scores in all domains significantly correlated with average categorization scores on a 5-alternative forced choice vocal emotion perception paradigm (Luo et al., 2018) and NCIQ scores from Physical- and Social- (but not Psychological-) functioning domains were positively correlated with average categorization scores on a 2-alternative forced choice paradigm (Von Eiff et al., 2022). To our knowledge, QoL has not been investigated directly for potential correlation with musical emotion perception. However, music appreciation has been significantly associated with NCIQ QoL scores (e.g., Fuller et al., 2021), which is in turn strongly linked to perceiving musical emotion (Garrido & Schubert, 2011; Grewe et al., 2005).

#### The current study

The current study aimed to provide a comprehensive account of auditory emotion perception in CI users. Firstly, we aimed to investigate whether the same CI user participants categorize vocal and musical emotions similarly and exhibit similar patterns of arousal and valence dimension perception across the two domains. Secondly, we aimed to explore potential associations between vocal and musical emotion categorization and individual differences in acoustic hearing, voice cue perception, as well as QoL. For vocal emotion categorization, we used the EmoHI test, previously used with CI children (Nagels et al., 2020), with three emotion categories of happy, angry, and sad (Experiment 1). For musical emotion categorization, we used a paradigm previously tested on NH participants with CI simulated classical music excerpts, with four emotion categories of joy, fear, serenity and sadness (Filipic et al., 2010; Harding et al., 2023; Lévêque et al., 2018) (Experiment 2). The influence of acoustic hearing was explored by comparing outcomes between unilaterally implanted CI users who reported to have acoustic hearing (bimodal group) or no acoustic hearing (unilateral group) in addition to electric hearing with the CI (Experiment 3a). For voice cue perception, we measured voice discrimination in just-noticeable differences (JNDs) for F0 and VTL (Gaudrain & Başkent, 2015, 2018) (Experiment 3b), and for QoL assessment, we used the NCIQ (Hinderink et al., 2000) (Experiment 3c). We hypothesized that (1) if emotion perception mechanisms are similar across materials, vocal and musical emotion categorization outcomes would be correlated, and arousal and valence dimensions would display similar patterns; (2) acoustic hearing would yield higher vocal and musical emotion categorization scores; (3) voice cue discrimination thresholds and QoL scores would correlate with vocal emotion categorization in line with previous findings (e.g., Chatterjee et al., 2023; Von Eiff et al., 2022), and following reports that vocal and musical emotion perception are similar in CI users (Paquette et al., 2018; Volkova et al., 2013), these individual differences would also be reflected in musical emotion categorization.

## General methods

### Participants

Twenty-eight CI users, all native speakers of Dutch, participated in all Experiments 1, 2, and 3. Participants’ ages and other demographic information are summarized in Table 1. Originally, 30 CI users had signed up for the study, but two participants dropped out before participating in any of the tests. Two CI users’ data in Experiment 3a could not be included in all analyses due to lack of convergence in the adaptive test.

**Table 1.**
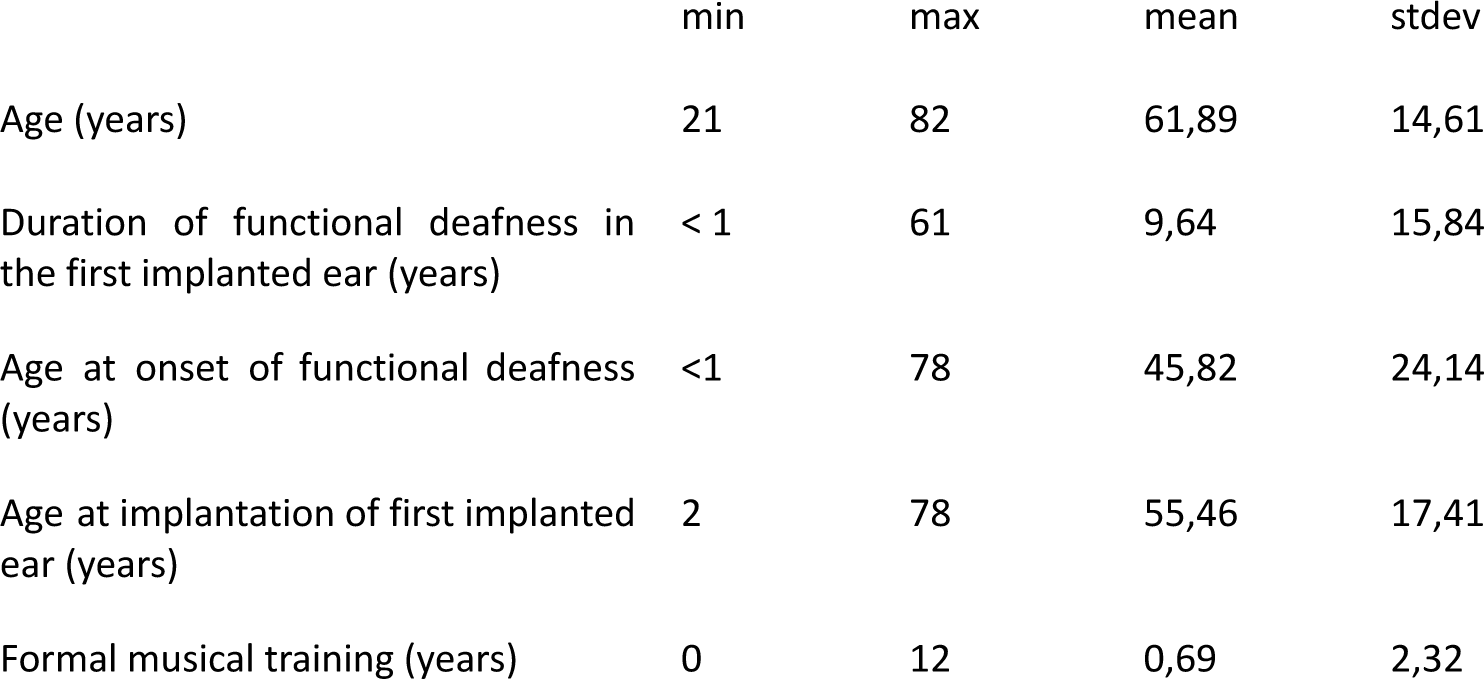
Summary of participant demographic information (*N* = 28). Functional deafness refers to non-functional hearing with optimized hearing aid(s) at the level of normal conversation.

|  | min | max | mean | stdev |
| --- | --- | --- | --- | --- |
| Age (years) | 21 | 82 | 61,89 | 14,61 |
| Duration of functional deafness in the first implanted ear (years) | < 1 | 61 | 9,64 | 15,84 |
| Age at onset of functional deafness (years) | <1 | 78 | 45,82 | 24,14 |
| Age at implantation of first implanted ear (years) | 2 | 78 | 55,46 | 17,41 |
| Formal musical training (years) | 0 | 12 | 0,69 | 2,32 |

Participants were recruited as part of a larger music training study (CIMUGAME) by letter, advertisement on social media, or word-of-mouth. As part of the inclusion criteria for the larger study, participants were older than 18 years, spoke native-level Dutch, did not have any language disorders or dyslexia, had (corrected) normal visual acuity, did not regularly engage in serious gaming, were healthy enough to participate in weekly piano or video lessons for six months, and had been using at least one implant for at least one year. There was no inclusion or exclusion based on CI configuration, as long as they had one cochlear implant, and regardless of pre- or postlingual deafness onset or early or adult implantation. All participants were residing in the Netherlands or Flemish Belgium. Implants from Cochlear (n = 20), Advanced Bionics (n = 3), and MED-EL (n = 5) were represented among participants. Further inclusion criteria were that the participants did not have more than three years of piano lessons and did not participate in any regular music making activities within the last three years of enrollment in the study, and were healthy enough to participate in weekly piano or video lessons for six months. Following these criteria, 22 participants had no formal musical training, with the remaining 6 as follows: (1) 12 years of flute lessons beginning from the age range 6-10 years old, (2) 2.5 years of piano lessons beginning from the age range 6-10 years old, (3) 2 years of organ lessons beginning from the age range 11-15 years old, (4) 1 year of piano lessons beginning from the age range 11-15 years old, (5) 1 year of piano lessons beginning from the age range 46-50 years old, (6) 3 months of voice lessons in adulthood (participant could not recall the exact year). The first 4 participants with instrument lessons had ended their formal musical training 44-45 years prior to enrollment in the study, the fifth participant ended lessons 20 years prior to enrollment, and none had been musically active in the interim years. The participant with voice lessons fulfilled the criteria of training being completed more than three years before the current study and was not involved in active music making before or since the voice lessons. This information is also summarized in Table 1.

Given the increasing number of bimodal CI users, and with research suggesting that low-frequency acoustic hearing may contribute to CI users’ perception of pitch, music, and music emotions, we did not make unimodal or unilateral CI use an inclusion/exclusion criterion. Instead, we opted to include CI users with or without acoustic hearing, with single or bilateral implants, to reflect the real-world variability of auditory perception in the general CI population. As a result of this non-restricted inclusion, we ended up with a relatively heterogeneous group in both demographic factors (Table 1) and in unilateral, bimodal, and bilateral CI use. Sixteen participants who were unilaterally implanted reported no acoustic hearing (unilateral group). Ten participants who were unilaterally implanted reported to have some functional acoustic hearing or use of hearing aids (bimodal group). Two participants had bilateral CIs. Details of unilateral and bimodal groups are presented in Experiment 3a.

All participants were informed about the study and provided written consent. The study protocol was approved by the Medical Ethics Committee of the University Medical Center Groningen (CIMUGAME, NL66549.042.20). Participants were reimbursed for travel costs to the testing location. As part of the larger study protocol, actual testing time was not reimbursed in monetary compensation, but was in exchange for receiving free piano or video game lessons.

### General procedures

As the present study was part of a larger musical training project (CIMUGAME), data were collected from the same population of CI adults in multiple experiments. All tests were conducted with the experimenter accompanying the participants and via an online-implementation using jspsych 6.1 software (de Leeuw, 2015), and data was automatically stored on a secure server. Data collection took place in a quiet setting, either in the lab or in a quiet room at the participant’s home or at the living facility. For Experiments 1, 2, and 3b, participants conducted the vocal and musical emotion categorization and voice cue discrimination tests on a Microsoft surface tablet using Logitech loudspeakers, placed approximately 10 cm to either side of the tablet. Participants were instructed to use their normal daily settings of the CI and, where applicable, also of the hearing aid. We did not plug the non- or second-implanted ear, nor turn off any devices, as part of the experiment. Before each test, participants adjusted the volume of their device(s) to a comfortable level during a loudness-adjustment prompt and were instructed not to adjust the volume for the entirety of each respective test. However, given the large test battery of the CIMUGAME project, as an exception, in the few occasions where participants reported stimuli to be too quiet for them to hear, we did allow an occasional volume adjustment. Screen backgrounds were dark gray or black with white text, where applicable. Response buttons were light gray buttons with dark gray or black labeling text. A progress bar showed how far along participants were during each test. For Experiments 3a-c, acoustic hearing was established by self-report from the participant and medical files later corroborated acoustic hearing thresholds. For Experiment 3c, QoL ratings were extracted from the NCIQ questionnaires.

### General analysis

All analyses were performed in R (R Core Team). Where applicable (across all experiments), multiple comparisons (post-hocs and sequential t-tests as well as correlations) were corrected using the false-discovery-rate method (Benjamini & Hochberg, 1995). All t-tests are the Welch variant for unequal variance and effect-sizes are reported using Cohen’s d. When ANOVAs (type III) were conducted, they were performed using the ‘ez’ package (Lawrence, 2016). The exact factors included in each ANOVA are described in the Results sections of each experiment. For each ANOVA, normality or sphericity was first tested to assess conformity with the assumptions required by the model. Effect-sizes are reported as generalized eta-squared (Bakeman, 2005).

## Experiment 1: Vocal emotion categorization

The aim of Experiment 1 was to assess vocal emotion perception in CI users. We used the vocal emotion test EmoHI, which previously has been used with child CI users and NH adults with vocoders (Nagels et al., 2020) and with children with hearing aids (Rachman et al., submitted). The EmoHI test uses basic emotion categories that fall in distinct arousal-valence quadrants to ensure task simplicity. Emotion categories are angry, happy, and sad, occupying three of the four arousal-valence quadrants (Figure 1; Table 2). A fourth stimuli category of neutral is also available as part of the EmoHI test, however, we did not include it here as neutrality by definition is towards the center of the valence and arousal continua, and would not contribute to assessing valence and arousal dimensions.

**Table 2.**
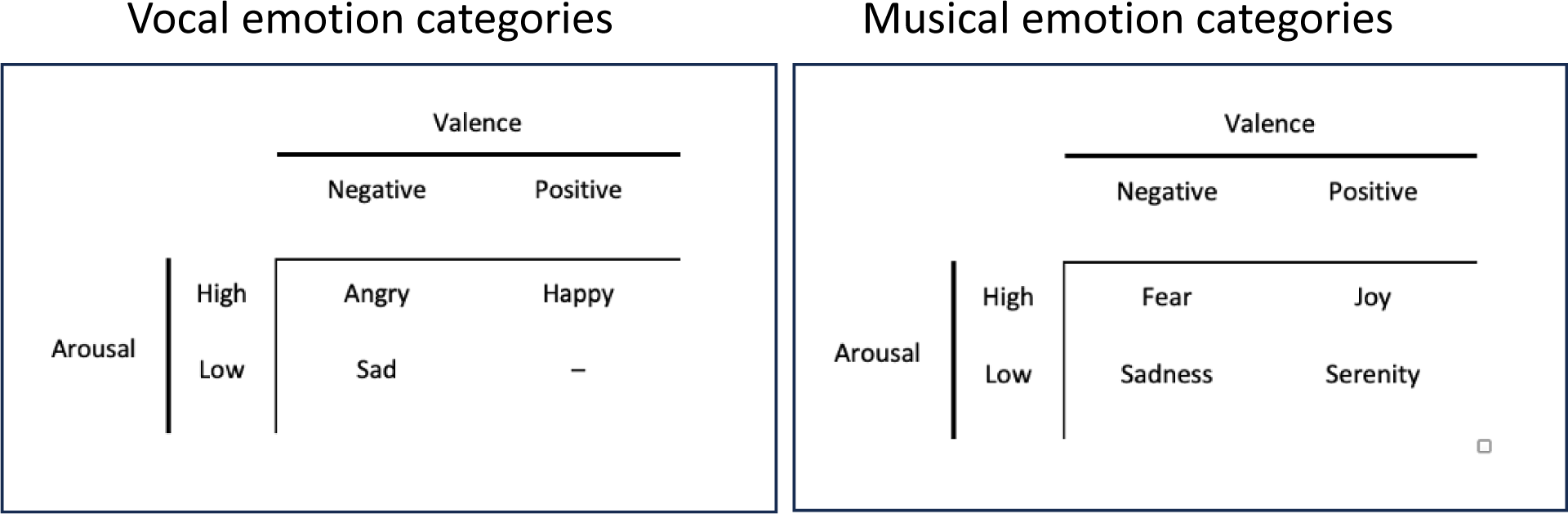
Vocal and musical emotion categories. Vocal emotion EmoHI test (left): Stimuli were 36 pseudo speech utterances exhibiting the emotion categories happy, angry, and sad. Musical emotion test (right): Stimuli were 40 instrumental classical music excerpts exhibiting the emotion categories joy, fear, serenity and sadness. In each test, each emotion category exhibited a unique combination of valence and arousal dimension characteristics, but the categories covered a different number of quadrants across the two tests.

While analysis of CI user’s sensitivity to individual emotion categories from the EmoHI has to our knowledge not yet been published, visual inspection has indicated that sensitivity to happy and angry was similar, while sensitivity to sadness was comparatively increased, in children with hearing aids (Rachman et al., submitted). Next to that, CI adults previously confused emotion categories happy and scared, both high-arousal but with opposing valence (Chatterjee et al., 2015; Lin et al., 2022). Taken together, we hypothesized in Experiment 1 that happy and angry would be confused because they are both high-arousal emotions.

### Materials and methods

#### Materials

The stimuli consisted of pseudospeech sentences ‘*Koun se mina lod belam’* and ‘*Nekal ibam soud molen’* that are not meaningful in any Indo-European languages and based on the Geneva Multimodal Emotion Portrayal (GEMEP) Corpus materials by Bänziger, Mortillaro & Scherer (2012). Materials were taken from the EmoHI vocal emotion perception test, described by Nagels et al. (2020). Stimuli were spoken by four native Dutch speakers (two male, two female) who were native monolingual speakers of Dutch without any discernible regional accent. Speakers were instructed to produce the sentences in a happy, sad, angry, or neutral (not used in this study) manner using emotional scripts that were also used for the GEMEP corpus stimuli (Bänziger & Scherer, 2010). The three emotions happy, angry, and sad were selected for the widest applicability across age ranges (Widen & Russell, 2004) and for maximum comparability of the EmoHI test across populations of differing hearing profiles (Nagels et al., 2020). Each speaker contributed three pseudo-speech sentences (two of one sentence, one of the other) for each emotion category. Stimuli were recorded in an anechoic room at a sampling rate of 44.1 kHz and equalized in RMS. Speakers provided written informed consent for the distribution and sharing of the recorded materials.

For practice, three additional stimuli (one per emotion, one production per speaker) were used for the training session, but not included in the experimental phase. For the experiment, 36 vocal emotion stimuli were used, comprising the categories happy, sad and angry, with 12 items in each category (4 speakers x 3 utterances). Each emotion category comprised both arousal and valence dimensions: either high or low arousal, or positive or negative valence (Table 2).

#### Procedure

Vocal emotion categorization was tested with the materials and methods of the EmoHI test (Nagels et al., 2020), with the difference that the current interface did not have a child-directed game-like environment. Participants were instructed to listen to the pseudo-sentences and determine whether the voice of the speaker sounded happy, sad, or angry, by clicking on one of three corresponding buttons on the screen labeled “blij”/happy, “verdrietig”/sad, “boos”/angry. Practice items, but not experimental items, were presented with feedback. Experiment items were presented in randomized order in one block of approximately 6 to 8 min.

#### Statistical analyses

The analysis techniques described in the ‘General analysis’ section above were applied to categorization sensitivity, d’, derived from raw categorization performance following the Signal Detection Theory (Green & Swets, 1988; Macmillan & Creelman, 2004) for each emotion category. For a given emotion category, the d’ was calculated by considering the correct categorization responses as hits (for a given row of the confusion matrix, the entry terms on the diagonal) and incorrect categorization responses, where the considered emotion was responded while not being presented, as false-alarms (the terms off the diagonal on the same row). The d’ was calculated as the difference between the z-transformed hit rate and false-alarm rate, divided by 2. If the hit or false-alarm rate was equal to 0 or 1, it was corrected by a quantity equivalent to a half trial to avoid infinite z-transformed values. If all trials were completed this corresponds to .04 and .96, respectively, yielding a maximum theoretical d’ of 2.45.

To further understand the type of error in categorization that participants made, the raw data of emotion categorization were compiled as confusion matrices. In a confusion matrix, a ‘perfect score’ would be represented as a diagonal line with points only on the same emotion category for both ‘presented’ (x-axis) and ‘responded’ (y-axis). Errors are represented by data points off the diagonal, or ‘confusions’, where a different category was responded from what was presented.

### Results

#### Vocal emotion categorization

Figure 2, panel A shows the d’ scores for vocal emotion categorization. On average across all participants, while all emotions yielded categorization significantly better than chance [*t*(27)>4.14, *p*_FDR_<0.001], the category ‘sad’ was best recognized (average d’=0.82), followed by ‘happy’ (d’=0.33), and ‘fear’ (d’=0.31).

**Figure 2.**
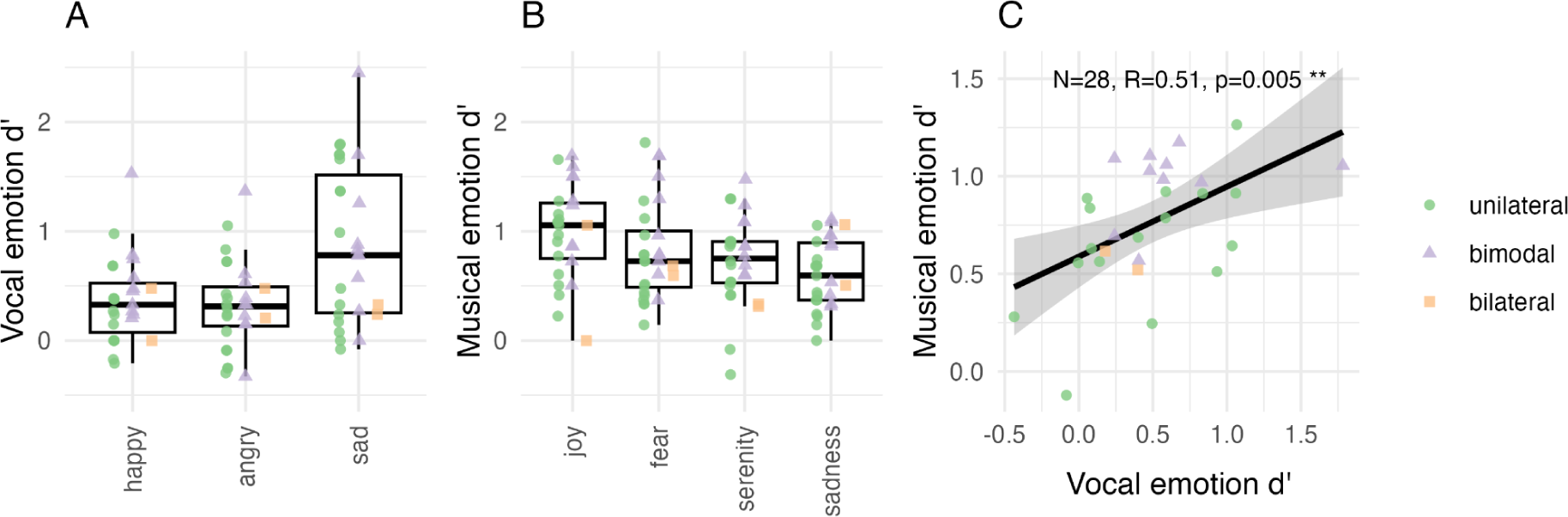
Emotion categorization scores shown in sensitivity (d’) for (A) vocal and (B) musical emotion perception, across all participants (*N* = 28). The horizontal line in the boxplot shows the median d’ across participants. The box extends from the 25^th^ to the 75^th^ percentile, and the whiskers extend to the value most remote from the median within 1.5 times the interquartile range. Individual data points are overlaid on top of the boxplots with unique shape and color for unilateral (green), bimodal (lilac), and bilateral (orange) implant users, respectively. d’ = 0 denotes the chance level. (C) Correlation of average d’ across emotion categories for vocal and musical emotion categorization, shown across all participants (*N* =28), The black line shows the regression along with the 95% confidence interval.

The d’ values were analyzed with a repeated-measure ANOVA with the factor Emotion category (‘happy’, ‘fear’, ‘sad’) as a repeated factor within participants. The ANOVA confirmed a significant main effect of emotion: sensitivity was different across presented emotion categories [*F*(2,54)=16.2, *p*<0.001, η^2^*_G_* =0.16]. Pairwise comparisons indicate that sensitivity for ‘sad’ categorization was significantly higher than for both ‘happy’ [*t*(27)=4.43, *p*_FDR_<0.001, *d*=0.84] and ‘angry’ [*t*(27)=4.52, *p*_FDR_<0.001, *d*=0.85], whereas the two did not significantly differ from each other [*t*(27)=0.22, *p*_FDR_=0.83, *d*=0.04].

##### Confusion matrices

Figure 3, panel A shows the confusion matrices with raw data for vocal emotion categorization, averaged across all participants (n = 28). Visual inspection of data indicates that confusion of emotion categories was more along the arousal dimension than the valence dimension. Emotion categories that shared an arousal class (happy and angry are both high arousal) were systematically confused, whereas the emotion category with a unique arousal class (sadness is low arousal) was seldom confused with other categories. This specific pattern of confusion is in line with the observation that “sad”, the only low arousal emotion category presented, yielded a significantly higher categorization sensitivity than the other two emotion categories, which are both high arousal.

**Figure 3.**
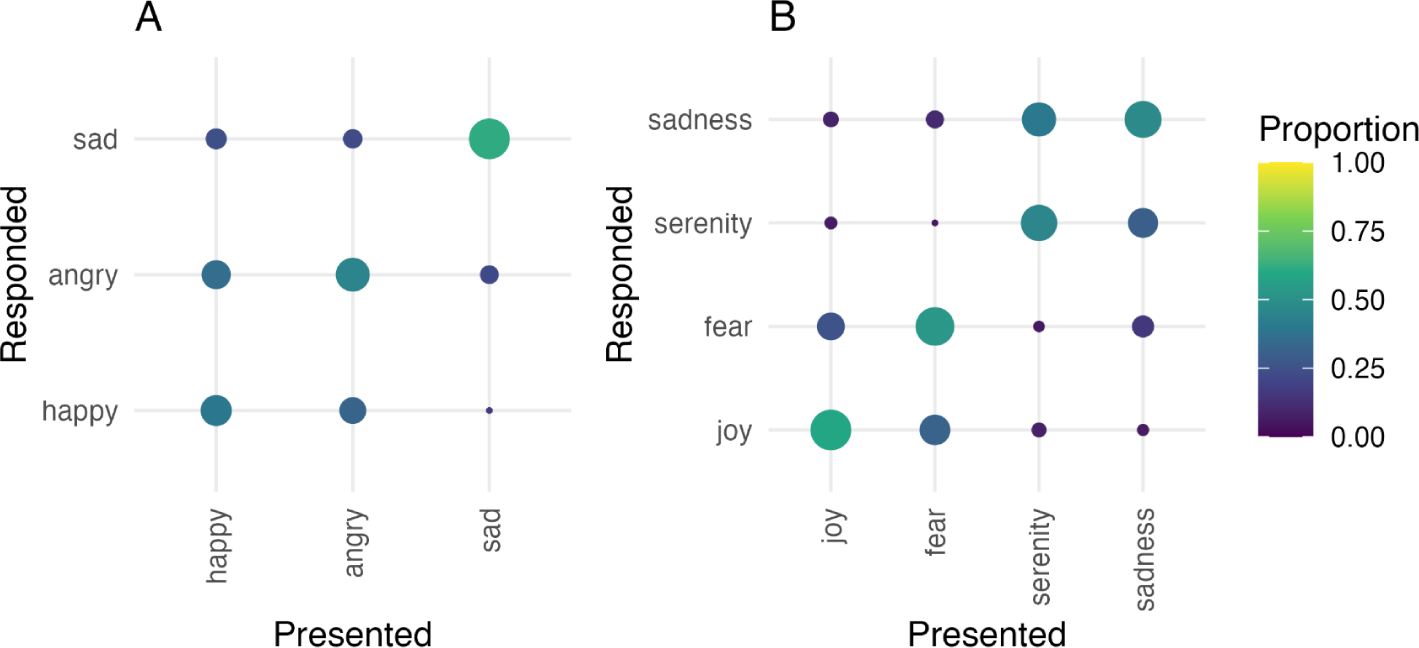
Confusion matrices shown for (A) vocal and (B) musical emotion categorization for all participants (*N* = 28). The size and color of each matrix entry is proportional to the number of relevant responses. The presented and responded emotion categories are listed on the x-axis and y-axis respectively.

### Discussion

Experiment 1 assessed vocal emotion perception in adult CI users. All vocal emotion categories, on average, were categorized above chance level. However, when individual scores were inspected, there were some at chance level for some participants, and overall, there was a large variation in categorization scores across individual participants.

Upon visual inspection, the d’ range in current adult CI user results is similar to the d’ range of CI children aged 4-16 who took the same EmoHI test (Nagels et al., 2020), a range that also overlapped with NH children but was below NH adults. This fits previous reports of vocal emotion perception in populations with hearing loss and hearing device users, namely that there is wide individual variation in vocal emotion categorization but generally with lower overall scores than NH peers (reviewed in Jiam et al., 2017).

Sadness was significantly better categorized than happy or angry. In a previous study using the EmoHI test with children with hearing aids, visual inspection of the emotion category values (assessed according to age) appeared to show the same pattern (Rachman et al., submitted). These results are in line with previous results showing that CI users confused high-arousal happy and scared emotion categories (Chatterjee et al., 2015; Lin et al., 2022). However, in emotion literature, sadness may have neural processing distinct from happy or angry (e.g., Harrison et al., 2007; Park et al., 2015), which could alternatively account for an increased sensitivity for sadness. This will be addressed in the general discussion. Experiment 2 investigated whether similar patterns of perception for vocal emotion were also found for musical emotion perception.

## Experiment 2: Musical emotion categorization

The aim of Experiment 2 was to assess musical emotion perception in adult CI users. We used the same musical excerpts and interface from our previous study with NH participants (Harding et al., 2023). These excerpts were taken from classical music pieces representing four emotion categories, joy, fear, serenity and sadness, which occupy all four quadrants in the valence and arousal dimensional space (Figure 1; Table 2) (Bigand et al., 2005).

When NH participants were tested with the full acoustic signal in Harding et al (2023), a main effect of emotion showed that for both full acoustic stimuli and vocoded stimuli, joy and fear had higher sensitivity than serenity and sadness. Thus we would expect a similar pattern of sensitivity for CI users. Inspection of arousal and valence dimensions in Harding et al. (2023) indicated that arousal and valence were both accurately perceived, arousal being perceived slightly better than valence. In contrast, when tested with vocoded excerpts (manipulating the temporal and spectral content available in musical excerpts to acoustically simulate spectrotemporal degradations), perception of arousal decreased slightly while perception of valence was at floor. Based on these vocoder simulation results, in Experiment 2 we hypothesized that CI users would perceive arousal significantly better than valence, if they could perceive valence at all.

### Materials and methods

#### Materials

Stimuli were 40 classical musical excerpts expressing the four emotion categories joy, fear, serenity, and sadness (10 items per category). Each category was of either high or low arousal, or positive or negative valence (Table 2). The original excerpts were all selected from recordings of orchestral arrangements containing multiple instruments and no vocals. The original stimuli were used by Bigand et al. (2005), Lévêque et al. (2018), Liégeois-Chauvel et al. (2014), Pralus et al. (2020), and Harding et al. (2023)^3^. The stimuli we used in this study were a subset of the stimuli reported by Harding et al (2023). Harding et al. (2023) indicated that the first and second halves of 20 s stimuli were similarly categorized in the emotions they expressed. Thus, 10 s excerpts only, randomly chosen from the first or second half of the original 20 s excerpt, were used to maintain shorter testing times in anticipation of testing with CI users. Especially as the test was part of a larger study and test battery, we wanted to prevent potential fatigue from long test durations.

Training items were 8 other items, not used in the experimental phase, which were comprised of two excerpts each of four works with iconic emotion status: Joy (Vivaldi’s *Four Seasons: Spring*) – Serenity (Monoman’s *Meditation*) – Fear (Mussorgsky’s *Night on Bald Mountain*) – Sadness (Beethoven’s *Moonlight Sonata*).

As described by Harding et al., (2023, supplementary material S4), loudness of the excerpts was adjusted, as the original recordings were obtained from different recordings, and hence had different loudness values that were not aligned with the intended loudness of the original piece. In normal hearing this is not an issue for testing because they can perceive acoustic details in the excerpts that inform them on *intended* loudness, i.e. how loud the musicians are playing as opposed to the physical intensity level of the recording. However, in CIs, these acoustic details are most likely to be lost, which could lead listeners to rely solely on physical loudness rather than intended loudness. Since intended loudness is a cue that is relevant for the expressed emotion of the musical piece, a mismatch between intended loudness and physical loudness can be a misleading cue for CI users. To account for this, a loudness adjustment was applied to re-align the physical loudness with the putative intended loudness estimated by listeners with musical expertise. Eleven musicians with different levels of musical expertise rated the softest and loudest parts of each piece by clicking buttons ranging from very soft (pianissimo) to very loud (fortissimo). These ratings were correlated with physical loudness computed with the Cambridge loudness model (Glasberg & Moore, 2002).

#### Procedure

Musical emotion categorization was tested with the same game-like interface and methods described by Harding et al. (2023), where participants were asked to ‘help’ aliens understand earth music. Participants were instructed to listen to a musical excerpt and to categorize which emotion was evoked by the music, by pressing one of four response buttons corresponding with the emotions “vreugde”/ joy, “droefheid” / sadness, “angst” / fear, “kalmte” / serenity. Training items were presented with feedback, but during the test, there was no feedback on correctness of categorization. Each emotion category was tested with 10 excerpts, resulting in a total of 40 trials per participant. Stimuli items were presented in random order per participant in two blocks of 20 items each, separated by a self-paced break.

After each categorization trial, a confidence in the categorization decision was assessed for maximum comparability with the paradigm of Harding et al. (2023). After each categorization response, participants were presented with an horizontal analog confidence scale, the leftmost position of the scale labeled 1 “Heel weinig vertrouwen”/(Very little confidence) and the rightmost position labeled 7 “Heel veel vertrouwen”/(A great deal of confidence). Tick marks were at integers from 1 to 7. Participants were asked “Met hoeveel vertrouwen heeft u deze categorisering gemaakt?”/(With how much confidence did you make this categorization?). A blue bar always started at 4, the midpoint, and participants had to drag the bar to the correct location for their answer. Any location on the scale was possible, i.e., not only at displayed numbers 1-7. Ratings were rescaled from 0 to 100 for analysis. Confidence ratings are reported in supplementary materials S1.

On average, the test lasted approximately 13 minutes, including the self-paced break. Participants took approximately 3 minutes for the break.

#### Statistical analyses

Like for the vocal emotion task in Experiment 1, the raw responses were converted to d’. Given the number of repetitions per emotion, the maximum theoretical d’ value is here 2.33. The other statistical methods are identical to those of Experiment 1.

### Results

#### Musical emotion categorization

Figure 2, panel B shows the d’ scores for musical emotion categorization. Mean sensitivity (*d*’) was significantly above zero, i.e., the scores were better than chance, in each emotion category (joy, fear, serenity, sadness) [*t’s*(28)>7.95, *p_FDR_’s*<0.0001], indicating that participants could carry out the task.

A repeated-measure ANOVA on individual d’ values with Emotion category as repeated factor within participants, showed that some emotions were better categorized than others [*F*(3,81)=5.05, *p* =.003, η^2^*_G_* =.084]. Comparing all emotions in pairs, only ‘sadness’ significantly differed from ‘joy’ [*t*(27)=3.71, p_FDR_<0.01, d=0.70] and from ‘fear’ [*t*(27)=2.62, *p_FDR_*<0.05, *d*=0.50], while all the other comparisons were not significant [*t*(27)<1.95, *p_FDR_*>0.12].

To assess within-participant perceptual similarities between vocal and musical emotion perception, musical emotion d’ was further entered into a Pearson’s product-moment correlation with vocal emotion d’ scores. d’ scores were strongly correlated across vocal and musical emotion categorization tasks (*R*=0.51, *p*=.005; Figure 2, panel C). Bilateral CI users were included in this correlation (*N* = 28).

#### Confusion matrices

Figure 3, Panel B shows the confusion matrices with raw data for musical emotion categorization, averaged across all participants (*N* = 28). A visual inspection indicates that confusion of emotion categories occurred primarily within the arousal dimension (joy was confused with fear and vice versa, and serenity was confused with sadness and vice versa).

### Discussion

Experiment 2 assessed musical emotion perception in adult CI users. All musical emotion categories, on average, were categorized above chance level, however, there was a large variability of d’ scores across individual participants, and for some emotion categories, some participants did show chance level scores.

The findings replicated the results with NH participants tested with vocoders in a previous study (Harding et al., 2023). The d’ range of current adult CI users was similar to the d’ range of vocoded excerpts reported by Harding et al. (2023) and below that of the d’ scores with the fully presented, non-vocoded acoustic signal.

With respect to categories, joy was categorized with the most sensitivity and sadness with the least. This is in line with the overall sensitivity reported with NH participants with the vocoded version of the current musical emotion stimuli (Harding et al., 2023). This finding generally aligns with visual inspection of accuracy scores in Ambert-Dahan et al (2015) (d’ and mean scores not reported), where CI users categorized musical emotion in classical excerpts with four emotions: happiness, fear, sadness and peacefulness. The result is also in line with Paquette et al. (2018), who reported joy to be the only emotion category identified above chance by CI users in short note bursts containing four emotion categories: joy, fear, neutral and sad. All in all, our sensitivity results demonstrate the viability of vocoder paradigms as a proxy of adult CI user musical emotion categorization, and show that the musical emotion categorization among current CI users is comparable to previous reports.

As indicated by the confusion matrix, the perception of musical excerpts as high (joy, fear) or low arousal (serenity, sadness) was easier than perceiving musical excerpts as positive (happy, serenity) or negative (fear, sadness). These findings are also in agreement with previously reported CI users’ categorization of happy/sad music emotions in piano melodies with chordal accompaniment, where their categorization relied on tempo cues but not on modal cues (Caldwell et al., 2015; D’Onofrio et al., 2020; Giannantonio et al., 2015; Hopyan et al., 2016). Thus, we can link findings from these previous studies beyond lab conditions to a four-emotion category paradigm with natural orchestral pieces exhibiting each emotion. Results indicate that CI users are better able to perceive how exciting or relaxing, but not necessarily how positive or negative, classical music is. This is a promising direction for CI users to explore their own general music listening pleasure, and encourages participation in music cultural activities, and to seek out composers and works with frequent changes between exciting and relaxing passages.

The low categorization score for the sad emotion observed with the musical emotion perception test stands in contrast to our observation within the vocal emotion perception test, Experiment 1, where the sad emotion categorization score was the highest. This is likely the result of the difference in experimental design and the difference in the number of emotion categories between the two tests, as they were developed by different research groups. This observation warrants caution for future studies where each emotion categorization score needs to be interpreted within the experimental design and the specific emotion categories included. However, despite the design differences, musical emotion categorization significantly correlated with vocal emotion categorization. This would suggest that at least for CI users, the auditory cues indicating a difference in emotion categories might be attributed to similar sensory or cognitive mechanisms, perhaps also to the increased similarity in cues due to similarly degraded spectrotemporal details. This is also in line with previous studies that reported similar arousal and valence ratings of vocal and musical emotion stimuli (Paquette et al., 2018) and the ability of CI children to categorize both vocal and musical emotion categories (Volkova et al., 2013). Moreover, this finding may also support the idea of robustness for CI populations for the two emotion perception tests used in this study.

## Experiment 3: Individual differences with respect to acoustic hearing, voice cue discrimination, and quality of life

### Experiment 3a Acoustic hearing

Cochlear implantation candidacy has been steadily evolving (Snel-Bongers et al., 2018). One characteristic of the changing criteria is that more acoustic hearing is permitted at the time of implantation, and increasingly both younger and older implantation is recommended. This has resulted in an increasing number of bimodal CI users.

Given that the presence of even very limited low-frequency acoustic hearing can contribute to increased perception of voice cues (Başkent et al., 2018), potentially relevant for vocal emotion perception, we assessed whether vocal emotion perception might be better in participants who self-reported having acoustic hearing.

Moreover, the presence of acoustic hearing has previously been shown to improve musical emotion categorization (D’Onofrio et al., 2020; Giannantonio et al., 2015) and music appreciation overall (Fata et al., 2009; Gfeller et al., 2006). We therefore also explored whether musical emotion perception scores might be higher in the bimodal CI user group than in the unilateral CI group.

#### Materials and Methods

##### Participants

Sixteen participants were unilaterally implanted and reported to have no acoustic hearing (unilateral group). Ten participants were bimodal implant users, i.e., they were also unilaterally implanted, but reported having some functional acoustic hearing or the use of hearing aids (bimodal group). The two bilateral CI users’ data were excluded as they did not fall in either category of unilateral or bimodal CI user. Tables 3 and 4 present the demographics factors for unilateral and bimodal groups separately. We did not include formal musical training in these tables since the range was already small, as was reported in Table 2.

**Table 3:** Summary of participant demographic information for the unilateral group (n = 16).

|  | min | max | mean | stdev |
| --- | --- | --- | --- | --- |
| Age (years) | 40 | 80 | 63,38 | 11,01 |
| Duration of functional deafness in the first implanted ear (years) | < 1 | 61 | 14,44 | 18,89 |
| Age at onset of functional deafness (years) | <1 | 77 | 40,88 | 25,50 |
| Age at implantation of first implanted ear (years) | 26 | 78 | 55,31 | 13,69 |

**Table 4:** Summary of participant demographic information for the bimodal group (n = 10).

|  | min | max | mean | stdev |
| --- | --- | --- | --- | --- |
| Age (years) | 28 | 82 | 63,70 | 15,64 |
| Duration of functional deafness in the first implanted ear (years) | < 1 | 22 | 3,60 | 7,82 |
| Age at onset of functional deafness (years) | 26 | 78 | 57,60 | 15,10 |
| Age at implantation of first implanted ear (years) | 26 | 78 | 61,20 | 15,58 |

Materials were the vocal (Experiment 1) and musical (Experiment 2) emotion tests.

#### Results

Figure 4 shows the vocal and musical emotion categorization scores (Panels A and B, respectively) for unilateral (n = 16) and bimodal (n = 10) CI users. The two bilateral CI users’ data were excluded as they did not fall in either category of unilateral or bimodal CI user.

**Figure 4.**
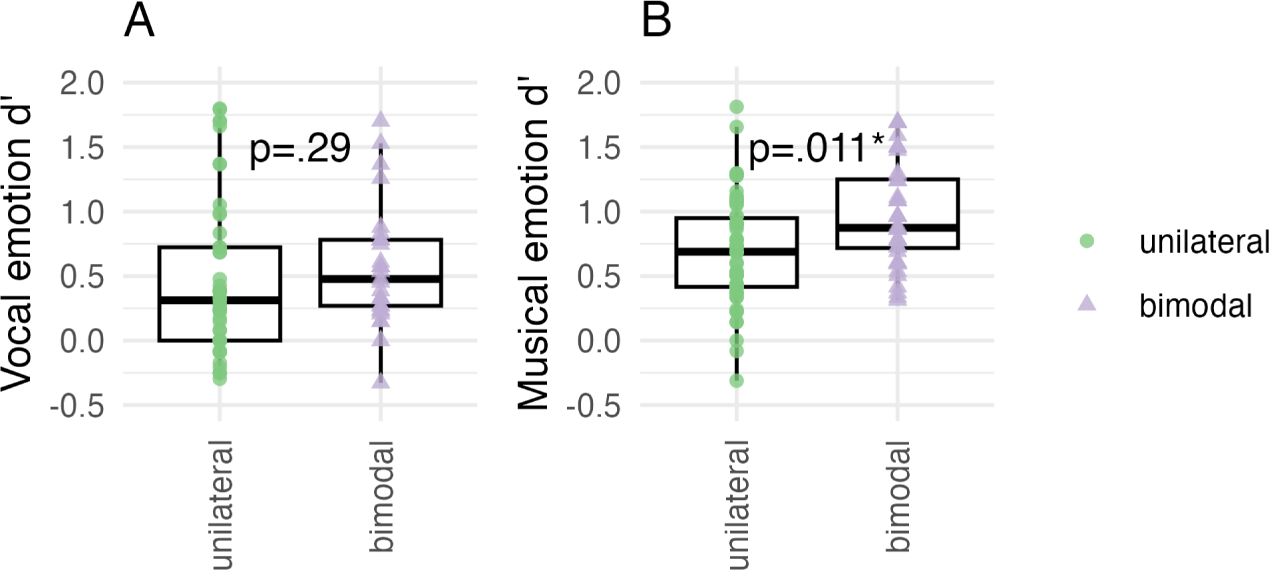
Emotion categorization d’ scores shown separately for the unilateral (green; n = 16) and bimodal (lilac; n = 10) participant groups for (A) vocal and (B) musical emotion perception. d’ scores from the bilateral (n = 2) implant user group are not included.

For vocal emotion categorization, a one-way ANOVA with factor Acoustic Hearing (unilateral, bimodal) compared overall vocal categorization sensitivity (d’) in unilateral and bimodal participants. The average d’ of the bimodal group was 0.63, against 0.43 for the unilateral group, however this difference was not significant (*F*(1,24), *p* =.290, η^2^*_G_* = .047).

For musical emotion categorization, a one-way ANOVA with factor Acoustic Hearing (unilateral, bimodal) compared overall musical categorization sensitivity (d’) in the two groups. The average d’ of the bimodal group was 0.97, against 0.66 for the unilateral group, and this difference was significant (*F*(1,24)=7.50, *p*=.011, η^2^*_G_* =.127).

#### Discussion

Contrary to our expectation, we did not observe a significant difference in vocal emotion perception between unilateral and bimodal CI users. This is at odds with previous findings indicating that the addition of low frequency acoustic hearing improves the perception of voice pitch (Başkent et al., 2018). This is assuming that voice cue discrimination would be improved in bimodal CI users, and that such improvement would be functionally relevant for the perception of emotion categories; however, this may indeed not be the case, a possibility that is followed up in Experiment 3b.

Overall musical categorization d’ was higher with the bimodal CI group than the unilateral CI group. This is in line with previous reports suggesting that the presence of acoustic hearing improves music perception in CI users (e.g., Fata et al., 2009).

### Experiment 3b Voice pitch and vocal tract length

When perceiving complex auditory input related to vocal and musical emotions, the extraction of specific auditory cues can be a limiting factor for CI users. In order to explore potential parallels between specific cue perception and vocal and musical emotion perception, we selected the voice cue discrimination metric from an available test battery for these participants that was tested for a larger training study (Başkent et al., 2018; Gaudrain & Başkent, 2015; Nagels et al., 2020). The voice cue discrimination test yields sensitivity thresholds, measured in JNDs for F0 and VTL. F0 and VTL perception have previously been linked to vocal emotion perception (Barrett et al., 2020; Deroche et al., 2016; Lin et al., 2022; Nussbaum et al., 2022; von Eiff et al., 2022). While we are not aware of any data showing that these acoustic voice cues are related to musical emotion perception, mean pitch in music is related to modal cues, which in turn inform valence features (Schubert, 2004). VTL, or in its more general form, acoustic scale, has been proposed as a perceptual dimension related to the notion of register in music (Patterson et al., 2010). Furthermore, VTL JNDs are thought to be an indicator of functional spectral resolution (Gaudrain & Başkent, 2015) and have therefore been suggested as a general indicator of implantation outcome (El Boghdady et al., 2019). They could therefore also be a predictor of musical emotion perception in as much as they are sensitive to the ability to perceive acoustic details. Therefore, Experiment 3b reports JNDs for voice cues of F0 and VTL and their correlations with vocal and musical emotion categorization. We also hypothesized that the presence of acoustic hearing would result in bimodal users having lower voice cue discrimination thresholds, and we therefore report JNDs of unilateral and bimodal CI users separately.

#### Materials and Methods

##### Materials

Stimuli used during the voice discrimination task consisted of three-syllable consonant-vowels (CVCVCV). Individual CVs were spliced from the items in the Nederlandse Vereniging voor Audiologie (NVA) corpus, which consist of meaningful CVC Dutch words (Bosmana & Smoorenburg, 1995) as reported by (Gaudrain & Başkent, 2015, 2018). One of the 61 extracted syllables used in the original study (Gaudrain and Başkent, 2015) was excluded because the sound quality was judged insufficient, resulting in a total of 60 CVs reasonably covering the Dutch phonological inventory. Average F0 and VTL were manipulated via analysis-resynthesis as implemented in the WORLD vocoder (Morise et al., 2016). The duration of the individual CV syllables originally varied between 142 and 200 ms, but they were all normalized to 200 ms during the WORLD resynthesis. To produce the CVCVCV triplet, for each trial, three individual CV syllables were randomly selected and concatenated with a separation of 50-ms of silence.

##### Procedure

The voice discrimination test was adapted from Gaudrain and Başkent (2015, 2018) and was identical to that used by Nagels et al. (2020), except for the child-friendly game-like graphical interface that was used with children and for the fact that no extra F0 contour was added on the triplet of syllables. JND measurements were acquired through an adaptive 3I-3AFC (three-interval, three-alternative forced choice) paradigm with a two-down, one-up rule (aiming at the 70.7%-threshold).

In each trial, participants heard 3 CVCVCV triplets, one of which (the odd-one-out) differed from the other two (the reference voice) in average F0 or VTL. The triplets were separated by a 500-ms silence. In total, the duration of 3 CVCVCV-triplets was approximately 3100 ms. CV-triplets presented during 1 trial were the same for all three intervals. Participants were presented with response buttons labeled “1”, “2”, or “3” while the three CVCVCV triplets played. The response button lit up during the playback of the corresponding triplet, with the target voice randomly placed among the first, second, or third triplet choice. The participants were instructed to click on the button that corresponded to the stimulus that was different from the two others. Once the participant made a choice, visual feedback was provided by the response button, turning green and bouncing vertically for correct responses, or red and shaking horizontally for incorrect ones.

The voice differences between the odd-one-out and the reference voice were produced by adjusting the F0 and VTL voice cue parameters using the WORLD vocoder as described above. The initial voice difference in each adaptive run was large, 12 st, in the direction of a male voice (the reference voice being, with a step size of 2 st. Step size adjustments occurred after every 15 trials or when the voice difference became smaller than twice the step size, dividing the step size by √2 to prevent negative differences. The maximum measurable difference was 25 semitones for both F0 and VTL dimensions. The adaptive run was repeated in a second block.

Each adaptive run ended after 8 reversals, with JNDs calculated from the last 6 reversals by taking their geometric average. If the number of trials exceeded the maximum number of trials of 150, or the voice difference became too large due to consecutive incorrect responses, the test was terminated, and its value was treated as missing data.

To acquaint participants with the task, four training trials (not part of the main task) were administered with large differences in F0 and VTL. The experiment comprised two five-minute blocks of stimuli separated by a self-paced break, and lasted about 10 minutes.

##### Statistical analyses

For two participants, the adaptive runs did not converge, leading to missing data values. This resulted in 26 participants (14 unilateral, 10 bimodal) with F0 values and 27 participants (15 unilateral, 10 bimodal) with VTL values. JNDs were log-transformed to create a normal distribution for statistics but VTL JNDs still did not pass a Shapiro-Wilk test of normality, therefore non-parametric tests were used for statistical analyses. When assessing the effect of acoustic hearing, two bilateral participants were excluded. Wilcoxon tests were used to compare JNDs across unilateral and bimodal participants, and Spearman correlations were used to assess associations between JNDs and vocal- and musical emotion d’ from Experiments 1 and 2.

#### Results

Figure 5, Panels A and D show the voice cue JNDs (st) for F0 and VTL. F0 JNDs were not significantly different between unilateral (n= 14) and bimodal (n = 10) groups (*W* = 76, *p* = .97). Similarly, VTL JNDs were not significantly different between unilateral (n= 15) and bimodal (n = 10) groups (W = 79, *p* = .63).

**Figure 5.**
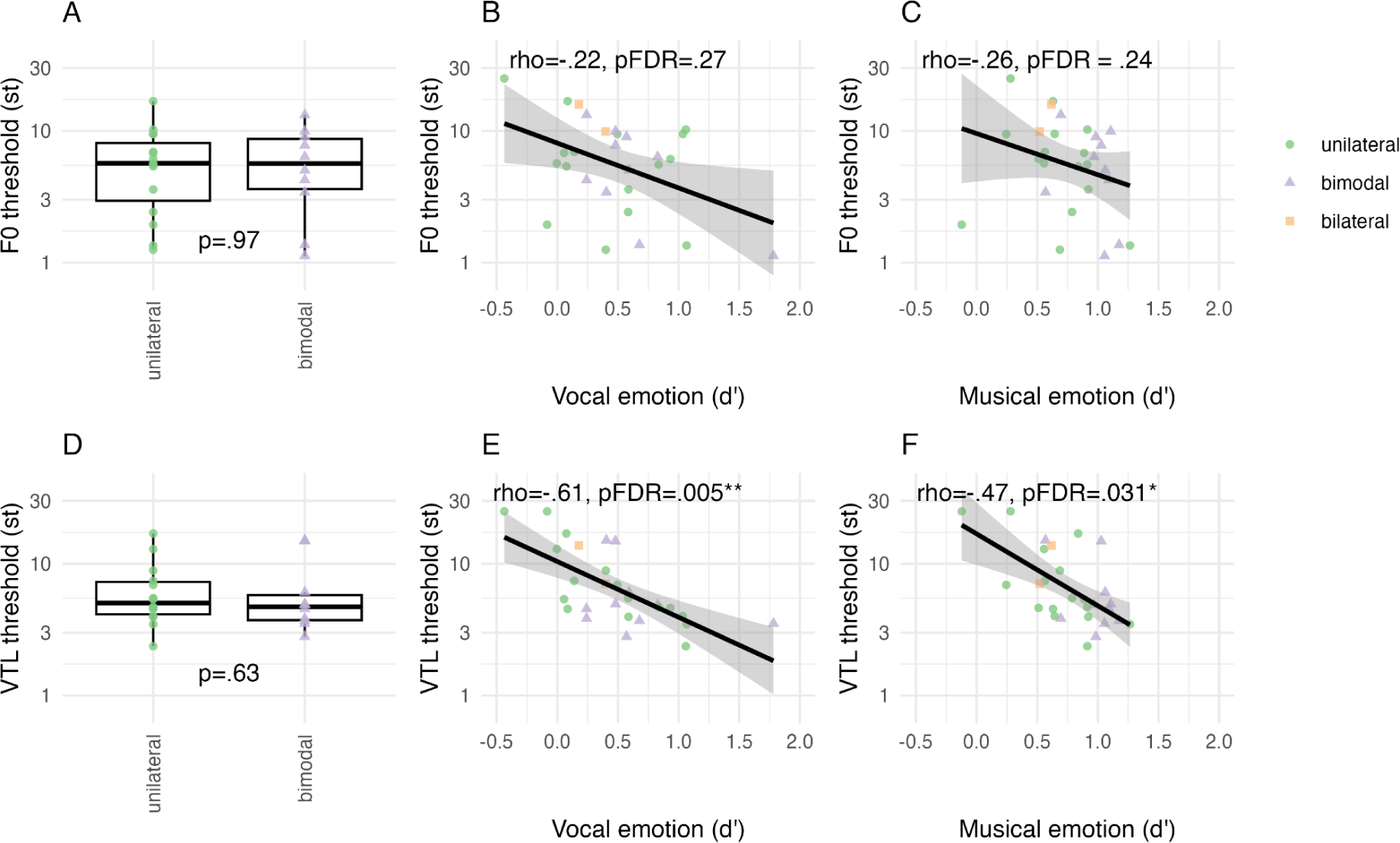
Voice cue discrimination JND thresholds for F0 and VTL shown separately for unilateral (green; n = 14 for F0, and n = 15 for VTL) and bimodal (lilac; n = 10 for both F0 and VTL) participant groups for (A) vocal and (D) musical emotion perception. JND data from n = 2 bilateral users were excluded. Correlations of JND scores with d’ scores shown for (B) F0 and vocal, (C) F0 and musical, (E) VTL and vocal, and (F) VTL and musical emotion categorization, shown across all participants (*N* = 26) whose JNDs converged. The black line shows the regression along with the 95% confidence interval.

Figure 5, Panels B,C,E, and F, show the correlations of the F0 and VTL JNDs with vocal- and musical emotion categorization d’ scores. F0 JNDs (n = 26) did not significantly correlate with vocal- (rho = -.22, *p* = .27) or musical emotion (rho = -.26, *p* = .24) d’ scores. VTL JNDs (n = 27) significantly correlated with both vocal- (rho = -.61, *p* = .005) and musical emotion (rho = -.47, *p* = .032) d’ scores.

#### Discussion

Voice cue JNDs in F0 and VTL were explored in Experiment 3b. The observed threshold ranges in adult CI users were comparable to ranges in vocoded thresholds with NH participants (e.g., Başkent et al., 2018; Biçer et al., 2023; Gaudrain & Başkent, 2015) and previously reported thresholds for adult CI users (e.g., Gaudrain & Başkent, 2018).

F0 thresholds did not correlate with vocal emotion categorization, or musical emotion categorization. This is in contrast to previous reports with CI users (Barrett et al., 2020; Deroche et al., 2016; Lin et al., 2022). Perhaps our CI users were more heterogeneously spread because we allowed bimodal users to use daily listening settings of their hearing aids as opposed to plugging their ear. However, Deroche et al. (2016) showed that children and adult CI users and NH participants all contributed to the correlation, obviously covering a large range of acoustic hearing in the NH population. Perhaps the large age-range of our study and the concentration of age in the older adult (60+) range may be causing more variance that prevents a significant correlation. While investigation of age effects is outside the scope of this study, it may be informative for future research that could systematically investigate F0 threshold and vocal emotion in younger vs older CI adults.

While the same argument for age applies to a link between F0 and musical emotion, there is another intuitive explanation why F0 perception did not correlate with musical emotion perception. Extracting the mean pitch from instrumental music with multiple instruments, such as in the current stimuli, may require extra processing steps to extract from multiple sources that the single-voice JND stimuli did not capture. Tools such as musical scene analysis (e.g., Hake et al., 2023) may be more appropriate for the investigation of potential correlations with musical emotion categorization.

VTL thresholds correlated with both vocal and musical emotion categorization. These results for vocal emotion categorization are in line with previous findings showing that combined periodicity, formant frequencies and spectrum level are strongly weighted when categorizing vocal emotion (von Eiff et al., 2022) in CI users who also weighted F0 as a cue to categorize emotions, as well as in CI users who did not weight F0 cues at all in their categorization choices. Interestingly, von Eiff et al. (2022) used different vocal emotion categories from our study, namely anger and fear. These emotion categories are both on the high-arousal, negative valence side of the emotion quadrant (see Figure 1), suggesting that the ability to perceive valence in vocal emotion may be linked to the ability to perceive these combined formant-related cues. Regarding musical emotion categorization, the observed correlation is in line with previous findings showing the use of spectral cues in the rating of arousal features in short musical bursts of notes (Paquette et al., 2018). Moreover, the VTL perception measure was previously speculated to be a general reflection of implant functioning (El Boghdady et al., 2019; Gaudrain & Başkent, 2018)). Our findings thus suggest that spectral cues that are also linked to VTL perception are salient to both vocal and musical emotion perception, and may also be a reflection of generally improved implant performance.

### Experiment 3c Quality of life

The NCIQ is a hearing-related QoL metric developed specifically for CI users (Hinderink et al., 2000). NCIQ assesses the subjective QoL in three general domains, with a total of six subdomains: Physical functioning (Sound perception basic, Sound perception advanced, Speech production), Psychological functioning (Self-esteem), and Social functioning (Activity, Social interaction). The NCIQ was previously positively associated with vocal emotion categorization in all subdomains, both with individual and total scores (Luo et al., 2018), and in all subdomains except for Self-esteem (Von Eiff et al., 2022).

Previous findings have shown a positive correlation between the total NCIQ score and a subdomain of a music appreciation questionnaire that assessed self-perceived quality in perceiving basic elements of music relating to pitch, rhythm, melody, and timbre (Fuller et al., 2021). While there is thus not a direct report of NCIQ and musical emotion correlation, perceiving musical emotion depends on a combination of these basic elements of music, and is also one component of music appreciation (Garrido & Schubert, 2011; Grewe et al., 2005). Thus an association between QoL and musical emotion categorization is plausible, in particular considering the link between QoL and vocal emotion from previous literature (Luo et al., 2018; von Eiff et al., 2022; Von Eiff et al., 2022) and the strong correlation between vocal and musical emotion categorization shown in Experiment 2.

Acoustic hearing may also influence QoL. For example, if low-frequency noises are perceptible with residual hearing, then hearing a bus or train rumbling by, or other basic sound perception features, may be more discernible. Acoustic hearing also involves a reported improvement in music appreciation (Fata et al., 2009; Gfeller et al., 2006), thus it might also influence how one perceives advanced complex sounds like music, or whether one chooses to engage in more activities like attending concerts.

Therefore, in Experiment 3c we assessed whether (1) the NCIQ ratings for QoL were associated with vocal or musical emotion categorization, by analyzing correlations with d’ for each subdomain, and (2) whether the presence of acoustic hearing influenced responses, by comparing the NCIQ QoL ratings between the unilateral and bimodal groups per subdomain.

#### Materials and Methods

##### Materials

The NCIQ (Hinderink et al., 2000) was originally developed to assess the subjective perception of QoL in CI users in 6 subdomains. (1) *Sound perception basic* items asked questions about how well participants perceived their ability to discern sounds such as cars approaching, people moving around them, and environmental noise identification and spatial localization. (2) *Sound perception advanced* items asked similarly phrased questions to basic sound perception about perceiving sound quality and localization, but regarding more complex signals like speech and music. (3) *Speech production* items asked participants to assess how well they felt that they were able to express themselves in different aspects of speech communication. (4) *Self-esteem* items asked participants to rate how confident or justified they felt in different situations where they might have to ask others to make adjustments to accommodate their hearing limitations. (5) *Activity* items asked participants about situations that they might avoid because of the hearing demands or inherent acoustic properties, such as grocery shopping. (6) *Social interaction* items asked participants to rate how their hearing affected daily conversations or social events.

Each item consisted of a question and a Likert-type rating scale to grade the response. Response options for items 1-55 were “Never, Sometimes, Regularly, Usually, Always, and Not applicable”. The scale was reversed in approximately half of the items, i.e., the question was formulated such that a positive response was “Never” as in “I never feel excluded” or that a positive response was “Always” as in “I always can make contact with others.” For items 56-60 response options were “No, Poor, Fair, Good, Quite well, and Not applicable,” referring mainly to production ability, where “No” was always negative and “Quite well” was always positive, e.g., in answering “I can hold a telephone conversation quite well.” An English translation of all items of the NCIQ can be found in the appendix of Hinderink et al. (2000).

##### Procedure

Participants received a paper questionnaire from the examiner, and were given an explanation and the opportunity to ask questions about the content or instructions. Some participants chose to fill out the questionnaire in the lab, and some others, alternatively, chose to fill out the questionnaire at home, then mailed it back to the clinic with a postage-paid return envelope. Filling in the questionnaire took approximately 10-15 minutes.

##### Statistical analysis

Likert scale scores were rescaled such that the most positive answer for each item was 100 and the most negative answer was 0, and averaged per subdomain, such that each participant received an average score per subdomain between 0 and 100 (Hinderink et al., 2000). Items left blank or answered “Not applicable” were omitted from analysis and thus did not contribute to subdomain average scores. Normal distribution of the scores for each subdomain was verified with a Shapiro-Wilk test.

#### Results

Figure 6A shows the NCIQ QoL subdomain ratings. To compare ratings across subdomains and between CI users with and without acoustic hearing, rating averages were entered into a mixed ANOVA with Subdomain (Sound perception basic, Sound perception advanced, Speech production, Self-esteem, Activity, and Social interaction) as repeated factor within participant, and Acoustic Hearing (unilateral, bimodal) as factor between participants. The data from one unilateral participant was removed because their QoL rating was missing in one subdomain. The two bilateral participants were also removed, bringing the number of participants to *N*=25. Ratings were significantly different across subdomains (*F*(5,120) = 20.03, *p* < .0001, η^2^*_G_* = 212) but did not differ according to whether acoustic hearing was present (*F*(1,23) = 1.36, *p* = .25, η^2^*_G_* = .04) nor did the two factors interact (*F*(5,120) = .33, *p* = .90, η^2^*_G_* = .004). Pairwise comparisons indicated that Speech production was rated higher than other categories, with significantly higher ratings than Sound perception basic and Sound perception advanced (p’s < .0001) and marginal higher ratings than Activity, Self-esteem, and Social interaction (*p*’s < .069). Otherwise Social interaction, Self-esteem and Activity were all rated higher than Sound perception advanced (*p*’s < .039).

**Figure 6.**
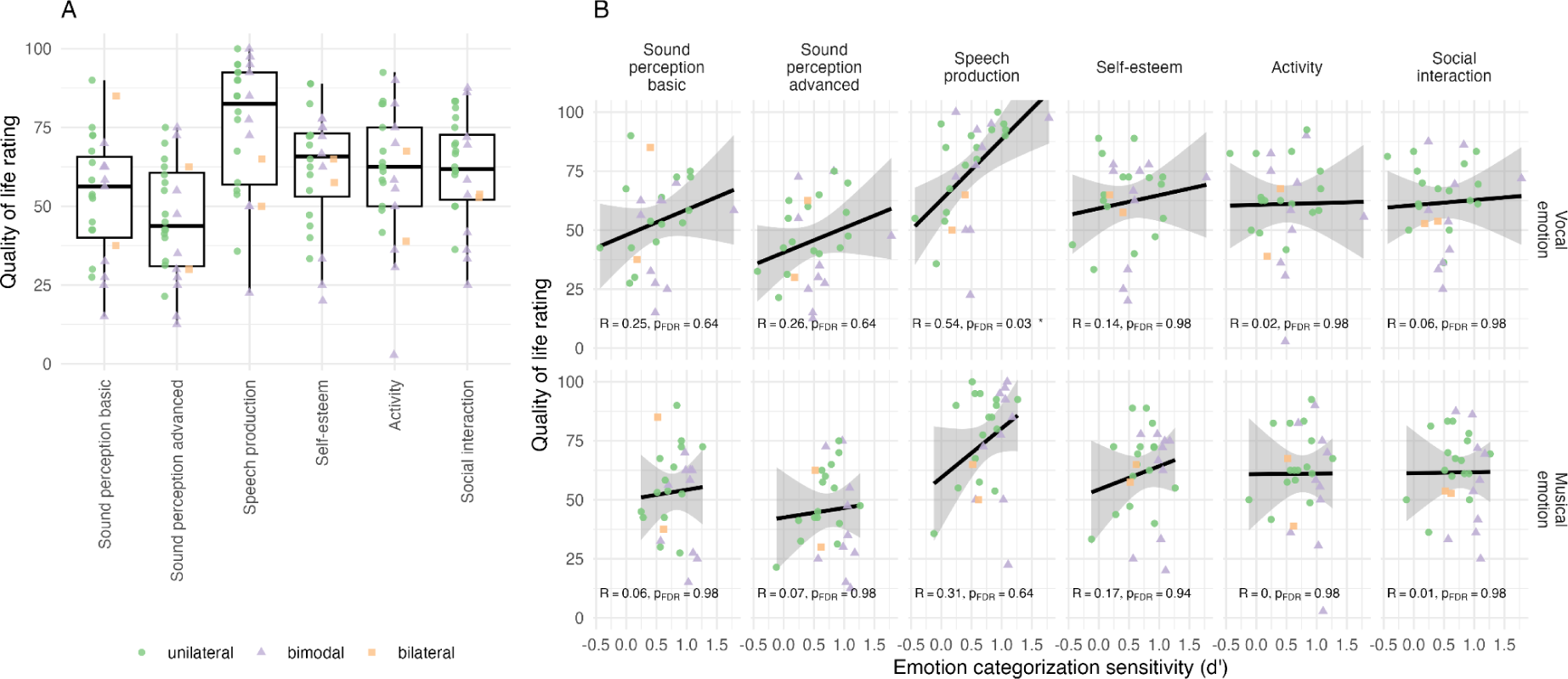
**(A)** Nijmegen Cochlear Implant Questionnaire subjective quality of life (QoL) ratings shown for the six subdomains and for all participants (*N*=25). **(B)** Correlations between the vocal (top) and musical (bottom) emotion categorization sensitivity (d’) and the QoL rating shown for each subdomain. The black line shows the regression along with the 95% confidence interval.

NCIQ subdomains were additionally entered into correlations with vocal and musical emotion sensitivity, with FDR-adjusted for 12 correlations (Figure 6B). Speech production scores correlated significantly with vocal emotion d’ (*R* = .54, *p* = .03), with no other significant correlation.

#### Discussion

QoL findings were comparable to previously published ranges for NCIQ QoL scores for CI adult users (Luo et al., 2018). The relatively high Speech production ratings reported by our participants may reflect the fact that a majority of our CI users (21 out of 28, based on self-reported hearing history) were postlingually deafened. It might also reflect a self-selection bias due to all participants being enrolled in a larger training study, i.e., participants who can communicate better with spoken language may be more willing to take on a long training challenge based on music. This same bias may also be at play regarding the relatively higher Self-esteem, Activity and Social interaction categories compared to Sound perception advanced; these participants are highly motivated CI users who lead active lives (including signing up for long training studies) despite their potential perceptual limitations, especially in advanced sound perception.

Previous studies with CI users showed that QoL subdomain scores significantly correlated with vocal emotion perception scores (Luo et al., 2018; von Eiff et al., 2022). In our study, not all subdomain measures significantly correlated with vocal emotion categorization scores, but the observed correlations and links with Speech production are in line with previous findings (Deroche et al., 2016; Barrett et al., 2020; Lin et al., 2022). One explanation for differences in the current results compared to previous studies where QoL-vocal emotion correlations were found could be that our group may have been more heterogeneous. Compared to Luo et al. (2018), we included both prelingual and postlingually-deafened CI users, as opposed to only postlingually deafened. Postlingually-deafened implant users can often recall QoL with functional acoustic hearing, which might make them rate QoL with the implant as lower. Moreover, compared to von Eiff et al. (2022), we included bimodal participants as opposed to participants who could only hear with one or two implants, i.e., eclectic hearing only. Thus, similar to the above argument, bimodal users with available acoustic hearing may have rated CI-related hearing lower in comparison to what they have available from the contralateral ear; indeed upon visual inspection of our data, bimodal users may have given slightly lower ratings, though not significantly different.

Finally, CI perception and production of vocal emotion have been linked previously in child CI users (Velde et al., 2019), supporting our finding that the QoL Speech production domain might exhibit the strongest association with vocal emotion perception. However, this does not match evidence from adult CI users (Chatterjee et al., 2019)his discrepancy will be addressed in the general discussion.

## General discussion

Across three experiments with adult CI users, the current study comprehensively investigated auditory emotion perception in both vocal and musical emotion perception, and whether individual differences in available acoustic hearing, voice cue discrimination and QoL were associated with emotion perception in either domain. We hypothesized that (1) if CI users’ emotion perception is similar across domains, categorizations in both domains would be correlated with each other and display similar confusion patterns along arousal and valence dimensions, and (2) individual differences in acoustic hearing, voice cue discrimination and QoL would be associated with vocal and musical emotion categorization. Supporting our hypothesis, we showed similarities between overall vocal and musical emotion perception scores and confusions for the emotion categories that fell in the same half-pane of the arousal dimension, indicating that the current categorization results are highly robust across CI users regardless of the test materials and designs. Partially supporting our hypothesis, we showed that cross-domain emotion perception taps into mechanisms linked to VTL perception, while domain differences emerge with respect to acoustic hearing, and QoL ratings.

### Vocal and musical emotion categorization with cochlear implants

Supporting our hypothesis, we found that vocal and musical emotion categorization displayed similar arousal and valence confusion patterns (depending on the specific design). This similarity occurred despite differences in the respective emotion tests. In the vocal emotion perception test, there were only three emotion categories, thus confusion only occurred for positive and negative valence and, high arousal emotions (happy, angry), and with high categorization accuracy for the single low-arousal emotion of sadness, since there was no confusion possibility with another emotion category. In the musical emotion perception test, confusion occurred between positive and negative valence for both high (joy, fear) and low (sadness, serenity) arousal emotions. These confusions are in line with previous findings, both for CI users and NH participants tested with vocoded stimuli (e.g., Chatterjee et al., 2015; Harding et al., 2023; Luo et al., 2007).

While similar valence confusion occurred for happy-angry in vocal emotion categorization and both joy-fear and sadness-serenity in musical emotion categorization, one large difference did emerge: Sadness was not confused with other categories in the vocal emotion test (in line with previous results with the same test; Rachman et al., submitted) whereas it was confused with another category, “serenity”, in the musical emotion test. On the one hand, this supports the interpretation that the arousal dimension is better perceived than the valence dimension in electric hearing with CIs, and underscores the observation that in categorization paradigms, sensitivity for any one category may depend on the other categories present. The argument that similarly located categories on arousal-dimension space reduce overall categorization accuracy previously motivated a choice to offer fewer emotion categories that cluster together in order to increase accuracy scores for CI users (Volkova et al., 2013). This line of thinking would suggest that categorization of sadness was simply improved for vocal emotion because no other low-arousal category was offered, in contrast to low-arousal serenity being available as a choice next to sadness for musical emotion.

On the other hand, correct categorization of emotions only partly relies on acoustic content. Perhaps not every emotion carries the same weight in how urgently they need to be recognized. For example, recognizing sadness in vocal emotion may have perceptual mechanisms that are distinct from recognizing joy or anger in vocal emotion. Sadness in facial expressions has been associated with qualitative differences in pupil size compared to fear, surprise or disgust (Harrison et al., 2007). Previously in a vocal emotion perception study, musicians had unique neural network activation in response to sadness (but not to anger, fear, or joy), which was attributed possibly due to musical training enhancing social communication skills compared to non-musicians (Park et al., 2015). Moreover, in Park et al. (2015), neural differences emerged when behavioral results did not emerge between categories, suggesting a dissociation between behavior and brain. This also implies that emotion processing (of sadness) goes beyond the acoustic cues available in the signal and involves more higher-order cognitive processes than other high arousal categories. Automatic responses to affective states are processed in ventral networks, whereas the cognitively effortful regulation of emotions is processed in dorsal networks (reviewed in Phillips et al., 2003), and evolutionarily speaking, high arousal is immediately informative in terms of signaling the need for physical response (i.e., to run from a predator, or that an angry person might strike out) while low arousal on the other hand is more social and cognitively relevant. We can thus speculate that perhaps high arousal of our happy or angry stimuli might trigger a “fight, flight or freeze” response and be processed more automatically, whereas processing low-arousal sadness may be more effortful and may tap into a different ventral network, resulting in a qualitatively different categorization pattern. However, since in the musical emotion categorization test sadness was confused with serenity, it does not seem that the high d’ scores for sadness categorization in the vocal emotion test were due to the specialness of this emotion, but rather were largely a result of the methodology. This could be explored in future studies using neuroimaging methods that are compatible with CIs (e.g., Saliba et al., 2016).

Despite sadness presenting less confusion for vocal emotion compared to musical emotion categorization, we found a strong correlation of overall categorization d’ scores across the two domains. Previously, cross-domain similarities were found with complex stimuli in child CI users (semantically neutral sentences and piano melodies with accompaniment; Volkova et al., 2013), and simple stimuli in adult CI users (short non-word exclamations and short note bursts; Paquette et al., 2018). Thus we show for the first time that complex stimuli in vocal and musical emotion perception paradigms are categorized similarly within adult CI users. Paquette and colleagues (2018) argued that by using short simple stimuli they could assess basic auditory emotion processing. Our present findings extend the similarity of vocal and musical emotion in primitive expressions to more natural listening contexts, notably of longer pseudo-speech utterances and instrumental music in adult CI users. Moreover, the strong correlation of scores across the two domains occurred despite domain differences in emotion categories and the number of category choices (as opposed to one-to-one matching of emotions in Volkova et al., 2013, and Paquette et al., 2018). This suggests the finding to be rather robust, as the differences in paradigms were ‘working against’ the similarity in findings.

We would like to stress that we cannot claim a common vocal-musical emotion processing mechanism that might generalize beyond hearing with CIs. Since the sounds delivered in electric hearing are lacking in spectrotemporal details, it could be that the speech and music auditory emotional cues resemble each other closely in electric hearing. In other words, emotional cues become similar across speech and music due to drastic cue reductions, where the only surviving cues are common across the domains.

### Individual differences and vocal and musical emotion perception

Partially supporting our hypothesis, we found that VTL discrimination, but not F0 discrimination, significantly correlated with both vocal and musical emotion categorization and that the QoL subdomain of Speech production significantly correlated with vocal emotion categorization, but not with musical emotion categorization.

F0 discrimination did not correlate with vocal emotion perception, as hypothesized and in line with previous literature (Barrett et al., 2020; Deroche et al., 2016; Lin et al., 2022). We suggested that perhaps some of our participants being of relatively older age in our CI population might play a role. Indeed compared to younger adults, older adults were reported to have lower accuracy in discrimination of F0 contour in both full acoustic (Zaltz & Kishon-Rabin, 2022) and simulated electric hearing degradations (Souza et al., 2011). Considering that older NH and CI adults have also reported lower accuracy in vocal emotion perception (e.g., Cannon & Chatterjee, 2022), perhaps the age-related perceptual changes for both metrics contributed to the association observed in younger NH and CI adults to become weaker.

We also explored whether F0 discrimination would correlate with musical emotion perception, considering that pitch interval perception is relevant to processing valence in music (Gomez & Danuser, 2007) and that valence in vocal and musical emotion was similarly rated in CI user adults (Paquette et al., 2018). However, F0 discrimination showed no statistically significant relationship with musical emotion perception. Thus, while age-related perception might play a role, a domain-specific metric for musical pitch intervals may be more relevant to musical emotion processing than voice F0, despite the cross-domain similarity of vocal and musical emotion sensitivity.

In contrast to F0, VTL discrimination correlated with both vocal and musical emotion perception. For vocal emotion, this supports previous results showing that both younger and older adults benefit from voice formant cues added to F0 cues when discriminating voice JNDs (Souza et al., 2011). However, it is, to the best of our knowledge, a novel finding that VTL JNDs correlate with musical emotion perception. On the one hand, this may be evidence that vocal and musical emotion perception mechanisms are similar across speech and music domains, if spectral information related to VTL is beneficial. On the other hand, this may be evidence that a VTL metric reflects a more general CI performance, as was suggested in a previous report (El Boghdady et al., 2019). We cannot distinguish between these two explanations. In the future, a battery of perceptual tests, in speech and music domains, as well as controlled environmental sounds, might be able to answer whether the VTL benefit is general to all auditory processing with the implant.

We note further that our selection of tasks took advantage of an available test battery from a larger training study. A more focused selection of vocal and musical emotion tests, and auditory cue discrimination tasks specific to the music-domain could provide more insight into the broader perceptual mechanisms that CI users may or may not be using during vocal and musical emotion feature perception.

We found that from all QoL ratings only the sub-domain of Speech production significantly correlated with vocal emotion categorization. This intuitively suggests that in speech communication, the better one can perceive the emotions of a speaking partner, the better one can match expressed emotions to meet the register of the speaking partner. This is in line with other findings with child CI users, who were not exposed to vocal emotion expression before CI hearing, linking their vocal emotion production with perception (Velde et al., 2019) reports that prelingually deafened children do not demonstrate as much acoustic variation in F0 production across emotion categories compared to their NH peers (Chatterjee et al., 2019). Using this argument for adult CI users does not align with reports that postlingually deafened adult CI users with no reported residual acoustic hearing, who heard vocal emotions before functional deafness and implantation, do not differ in the production of their acoustic features across vocal emotions (Chatterjee et al., 2019). However, our adult population includes both prelingually- and postlingually deafened CI users with and without acoustic hearing, who as a group may not resemble previous reports due to large heterogeneity.

Differences from previous adult CI user QoL correlations with vocal emotion — which included most or all subdomains from the NCIQ (Luo et al., 2018; von Eiff et al., 2022) — may be due to our participant sample reflecting a more contemporary and changing CI population. The first generation of prelingually-deafened, early-implanted children is growing up, and more adults and older adults are receiving implants with more relaxed criteria, for example, allowing more acoustic hearing in the contralateral ear and admitting increasingly older candidates, as well as a swifter transition in the audiology treatment plan from the stage where a hearing aid loses its function to afford speech comprehension in normal conversation (functional deafness) to implantation (Snel-Bongers et al., 2018). This reflects the heterogeneity previously reported in CI findings (Nyirjesy et al., 2023) and is thus a good representation of current CI users. In our case, this heterogeneity may have been advantageous in that a broad range in scores allowed us to capture individual differences in VTL and F0 as well as the correlation between the vocal and musical categorization.

### Acoustic hearing

Approximately one third of the participants self-reported (residual) acoustic hearing in their non-implanted ear that either was or was not amplified with a hearing aid. In Experiment 3a, there was no difference in the vocal emotion categorization between unilateral and bimodal CI users while there was a significant effect of acoustic hearing for musical emotion categorization. Moreover, in Experiment 3b, neither JNDs nor QoL for basic and advanced sound perception significantly differed between the unilateral and bimodal groups. One possible explanation might be that residual hearing was very limited, to the extent that it did not help discriminate pitch (in contrast to previous results with a CI simulation; Başkent et al., 2018). This is in line with recent results that showed no group differences between unilateral and bimodal CI users on a listening effort test battery (Nyirjesy et al., 2023), but that acoustic hearing still assisted in the detection of loudness changes. CIs reduce the dynamic range of transduced signals (Macherey & Carlyon, 2014), therefore bimodal users may have benefited more from the dynamic range of acoustic hearing. In musical excerpts, loudness changes were intentionally added to better distinguish emotion categories (Harding et al., 2023) while no loudness adjustment was made to the vocal emotion materials.

For the significant difference in musical emotion categorization, we inspected Tables 3 and 4 for the demographics factors, to rule out that some of these factors may have played a role. The average age and age range, as well as age at implantation of the unilateral and bimodal groups were almost identical. The onset of functional deafness was younger in the unilateral group than the bimodal group, which also resulted in longer duration of deafness in the unilateral group than the bilateral group. This difference possibly came from the fact that CI candidates with acoustic hearing did not really have a strong onset of deafness that they could identify since they always had some hearing. Duration of deafness was shown to be a factor that can negatively affect CI outcomes in some studies (Blamey et al., 2012; Plant et al., 2016), however, other studies cautioned that there would be more factors that would have a collective effect (e.g., Holden et al., 2013; van Dijk et al., 1999). Having said that, the acoustic hearing effect was only observed for musical emotion categorization, and not for vocal emotion categorization, which would be expected to be similarly affected by the same demographic factors. Hence, this point remains inconclusive from the current data. In future research, acoustic hearing thresholds and dynamic range per participant could be better characterized, if possible controlled for as an experimental variable, and incorporated into a balanced design for a more definitive answer.

## Conclusion

We showed that vocal and musical emotion categorization varied across the CI user participants, but the overall scores were strongly correlated across the two tests within the participants, despite the stimulus and design differences. Perhaps this is a consequence of the test population that was relatively heterogeneous, varying in age, pre vs. postlingual deafness and implantation, and acoustic hearing contribution. Despite this heterogeneity, for both vocal and musical emotion categorization, CI users’ relative perception of the arousal dimension seemed better than the valence dimension. Future research is needed to explore the relative contributions of sensory and neurocognitive mechanisms to explain these patterns. While age-related changes may have obscured some statistical associations with F0 discrimination, the emergence of VTL discrimination as correlating with both vocal and musical emotion perception lends weight to the hypothesis that VTL may be a proxy for general implantation outcome, warranting further investigation. Finally, considering that the vocal emotion test correlated with the musical emotion test, VTL discrimination, and QoL in terms of capability to communicate in Speech production, adding a vocal emotion test to a clinical assessment battery may aid clinicians to estimate a broader reaching impact of CI performance.

## Acknowledgements

We would like to thank Dr. Christina Fuller for support in conceptual planning of the study and proofreading the manuscript, Lisa Hoogeveen, Mirthe Ronde, and Isa Poortman for help with data collection, and Henk Mellema, Wijnna Beuving, Gesina Posthumus-Ottema, Esther Steenbergen-Bethlehem, and Rieta Steeman-Poelman for administrative support.

## Supporting information

Confidence ratings are reported in supplementary materials S1.

## Data Availability

All data that support the findings of this study will be openly available in the DataverseNL repository via (link forthcoming).

## Footnotes

1 Across musical cultures, valence associations with the same musical pieces can vary, while arousal is typically universally associated (Wang et al., 2022).

2 Visual inspection of results suggests clear category separation along dimensions, though rating values were not reported.

3 A full list of the stimuli material can be found in supplementary table S3 of Pralus et al (2020): https://doi.org/10.1016/j.cortex.2020.05.015.

## References

Ambert-Dahan, E., Giraud, A. L., Sterkers, O., & Samson, S. (2015). Judgment of musical emotions after cochlear implantation in adults with progressive deafness. Frontiers in Psychology, 6(MAR), 1–11. 10.3389/fpsyg.2015.00181

Bachorowski, J.-A., & Owren, M. J. (1995). Vocal expression of emotion: Acoustic properties of speech are associated with emotional intensity and context. Psychological Science, 6.

Bakeman, R. (2005). Recommended effect size statistics for repeated measures designs. Behavior Research Methods, 37(3), 379–384. 10.3758/BF03192707

Balkwill, L.-L., & Thompson, W. F. (1999). A Cross-Cultural Investigation of the Perception of Emotion in Music: Psychophysical and Cultural Cues. Music Perception, 17(1), 43–64. 10.2307/40285811

Banse, R., & Scherer, K. R. (1996). Acoustic profiles in vocal emotion expression. Journal of Personality and Social Psychology, 70(3), 614–636.

Bänziger, T., Mortillaro, M., & Scherer, K. R. (2012). Introducing the Geneva Multimodal expression corpus for experimental research on emotion perception. Emotion, 12(5), 1161–1179. 10.1037/a0025827

Bänziger, T., & Scherer, K. R. (2010). Introducing the Geneva Multimodal Emotion Portrayal (GEMEP) Corpus. In A blueprint for affective computing: A Sourcebook and manual. Oxford University Press.

Barrett, K. C., Chatterjee, M., Caldwell, M. T., Deroche, M. L. D., Jiradejvong, P., Kulkarni, A. M., & Limb, C. J. (2020). Perception of Child-Directed Versus Adult-Directed Emotional Speech in Pediatric Cochlear Implant Users. Ear & Hearing, 41(5), 1372–1382. 10.1097/AUD.0000000000000862

Başkent, D., Gaudrain, E., Tamati, T., & Wagner, A. E. (2016). Perception and psychoacoustics of speech in cochlear implant users. In A. T. Cacace, E. de Kleine, A. G. Holt, & P. van Dijk (Red.), Scientific Foundations of Audiology: Perspectives from Physics, Biology, Modeling, and Medicine. Plural Publishing.

Başkent, D., Luckmann, A., Ceha, J., Gaudrain, E., & Tamati, T. N. (2018). The discrimination of voice cues in simulations of bimodal electro-acoustic cochlear-implant hearing. The Journal of the Acoustical Society of America, 143(4), EL292-EL297. 10.1121/1.5034171

Belin, P., Fillion-Bilodeau, S., & Gosselin, F. (2008). The Montreal Affective Voices: A validated set of nonverbal affect bursts for research on auditory affective processing. Behavior Research Methods, 40(2), 531–539. 10.3758/BRM.40.2.531

Benjamini, Y., & Hochberg, Y. (1995). Controlling the False Discovery Rate: A Practical and Powerful Approach to Multiple Testing. Journal of the Royal Statistical Society: Series B (Methodological), 57(1), 289–300. 10.1111/j.2517-6161.1995.tb02031.x

Berg, R., & Stork, D. (2004). The Physics of Sound, 3rd Edition (3rd edition). Pearson.

Biçer, A., Koelewijn, T., & Başkent, D. (2023). Short Implicit Voice Training Affects Listening Effort During a Voice Cue Sensitivity Task With Vocoder-Degraded Speech. Ear and Hearing, 44(4), 900–916. 10.1097/AUD.0000000000001335

Bigand, E., Parncutt, R., & Lerdahl, F. (1996). Perception of musical tension in short chord sequences: The influence of harmonic function, sensory dissonance, horizontal motion, and musical training. Perception & Psychophysics, 58(1), 125–141. 10.3758/BF03205482

Bigand, E., Vieillard, S., Madurell, F., Marozeau, J., & Dacquet, A. (2005). Multidimensional scaling of emotional responses to music: The effect of musical expertise and of the duration of the excerpts. Cognition & Emotion, 19(8), 1113–1139. 10.1080/02699930500204250

Blamey, P., Artieres, F., Başkent, D., Bergeron, F., Beynon, A., Burke, E., Dillier, N., Dowell, R., Fraysse, B., Gallégo, S., Govaerts, P. J., Green, K., Huber, A. M., Kleine-Punte, A., Maat, B., Marx, M., Mawman, D., Mosnier, I., O’Connor, A. F., … Lazard, D. S. (2012). Factors affecting auditory performance of postlinguistically deaf adults using cochlear implants: An update with 2251 patients. Audiology and Neurotology, 18(1), 36–47. 10.1159/000343189

Bosmana, A. J., & Smoorenburg, G. F. (1995). Intelligibility of Dutch CVC Syllables and Sentences for Listeners with Normal Hearing and with Three Types of Hearing Impairment. Audiology, 34(5), 260–284. 10.3109/00206099509071918

Caldwell, M. T., Jiam, N. T., & Limb, C. J. (2017). Assessment and improvement of sound quality in cochlear implant users. Laryngoscope investigative otolaryngology, 2(3), 119–124. 10.1002/lio2.71

Caldwell, M. T., Jiradejvong, P., & Limb, C. J. (2016). Impaired Perception of Sensory Consonance and Dissonance in Cochlear Implant Users. Otology & Neurotology, 37(3), 229–234. 10.1097/MAO.0000000000000960

Caldwell, M. T., Ranklin, S. K., Jiradejvong, P., Carver, C., & Limb, C. J. (2015). Cochlear implant users rely on tempo rather than on pitch information during perception of musical emotion. Cochlear Implants International, 16(3), S114–S120.

Cannon, S. A., & Chatterjee, M. (2022). Age-Related Changes in Voice Emotion Recognition by Postlingually Deafened Listeners With Cochlear Implants. Ear and Hearing, 43(2), 323. 10.1097/AUD.0000000000001095

Chatterjee, M., Gajre, S., Kulkarni, A. M., Barrett, K. C., & Limb, C. J. (2023). Predictors of Emotional Prosody Identification by School-Age Children With Cochlear Implants and Their Peers With Normal Hearing. Ear & Hearing. 10.1097/AUD.0000000000001436

Chatterjee, M., Kulkarni, A. M., Siddiqui, R. M., Christensen, J. A., Hozan, M., Sis, J. L., & Damm, S. A. (2019). Acoustics of Emotional Prosody Produced by Prelingually Deaf Children With Cochlear Implants. Frontiers in Psychology, 10, 2190. 10.3389/fpsyg.2019.02190

Chatterjee, M., Zion, D. J., Deroche, M. L., Burianek, B. A., Limb, C. J., Goren, A. P., Kulkarni, A. M., & Christensen, J. A. (2015). Voice emotion recognition by cochlear-implanted children and their normally-hearing peers. Hearing Research, 322, 151–162. 10.1016/j.heares.2014.10.003

Chuenwattanapranithi, S., Xu, Y., Thipakorn, B., & Maneewongvatana, S. (2008). Encoding emotions in speech with the size code. A perceptual investigation. Phonetica, 65(4), 210–230. 10.1159/000192793

de Leeuw, J. R. (2015). jsPsych: A JavaScript library for creating behavioral experiments in a Web browser. Behavior Research Methods, 47(1), 1–12. 10.3758/s13428-014-0458-y

Deroche, M. L. D., Kulkarni, A. M., Christensen, J. A., Limb, C. J., & Chatterjee, M. (2016). Deficits in the Sensitivity to Pitch Sweeps by School-Aged Children Wearing Cochlear Implants. Frontiers in Neuroscience, 10. 10.3389/fnins.2016.00073

Deroche, M. L. D., Lu, H.-P., Limb, C. J., Lin, Y.-S., & Chatterjee, M. (2014). Deficits in the pitch sensitivity of cochlear-implanted children speaking English or Mandarin. Frontiers in Neuroscience, 8. https://www.frontiersin.org/journals/neuroscience/articles/10.3389/fnins.2014.00282

D’Onofrio, K. L., Caldwell, M., Limb, C., Smith, S., Kessler, D. M., & Gifford, R. H. (2020). Musical Emotion Perception in Bimodal Patients: Relative Weighting of Musical Mode and Tempo Cues. Frontiers in Neuroscience, 14, 114. 10.3389/fnins.2020.00114

Dorman, M. F., Gifford, R. H., Spahr, A. J., & McKarns, S. A. (2008). The Benefits of Combining Acoustic and Electric Stimulation for the Recognition of Speech, Voice and Melodies. Audiology & neuro-otology, 13(2), 105–112. 10.1159/000111782

El Boghdady, N., Gaudrain, E., & Başkent, D. (2019). Does good perception of vocal characteristics relate to better speech-on-speech intelligibility for cochlear implant users? The Journal of the Acoustical Society of America, 145(1), 417–439. 10.1121/1.5087693

Fata, F. E., James, C. J., Laborde, M.-L., & Fraysse, B. (2009). How Much Residual Hearing Is ‘Useful’ for Music Perception with Cochlear Implants? Audiology and Neurotology, 14(Suppl. 1), 14–21. 10.1159/000206491

Feldman, L. A. (1995). Valence focus and arousal focus: Individual differences in the structure of affective experience. Journal of Personality and Social Psychology, 69(1), 153. 10.1037/0022-3514.69.1.153

Filipic, S., Tillmann, B., & Bigand, E. (2010). Judging familiarity and emotion from very brief musical excerpts. Psychonomic Bulletin & Review, 17(3), 335–341. 10.3758/PBR.17.3.335

Fitch, W. T., & Giedd, J. (1999). Morphology and development of the human vocal tract: A study using magnetic resonance imaging. The Journal of the Acoustical Society of America, 106(3), 1511–1522. 10.1121/1.427148

Friesen, L., Shannon, R. V., Başkent, D., & Wang, Z. (2001). Speech recognition in noise as a function of the number of spectral channels: Comparison of acoustic hearing and cochlear implants. Acoustical Society of America, 110(2), 1150–1163. 10.1121/1.1381538

Fuller, C. D., Galvin, J. J., Maat, B., Başkent, D., & Free, R. H. (2018). Comparison of Two Music Training Approaches on Music and Speech Perception in Cochlear Implant Users. Trends in Hearing, 22, 1–22. 10.1177/2331216518765379

Fuller, C., Free, R., Maat, B., & Başkent, D. (2021). Self-reported music perception is related to quality of life and self-reported hearing abilities in cochlear implant users. Cochlear Implants International, 23(1), 1–10. 10.1080/14670100.2021.1948716

Garrido, S., & Schubert, E. (2011). Individual Differences in the Enjoyment of Negative Emotion in Music: A Literature Review and Experiment. Music Perception, 28 why do(3), 279-296.

Gaudrain, E., & Başkent, D. (2015). Factors limiting vocal-tract length discrimination in cochlear implant simulations. The Journal of the Acoustical Society of America, 137(3), 1298–1308. 10.1121/1.4908235

Gaudrain, E., & Başkent, D. (2018). Discrimination of Voice Pitch and Vocal-Tract Length in Cochlear Implant Users. Ear and Hearing, 39(2), 226–237. 10.1097/AUD.0000000000000480

Gelder, B., Teunisse, J.-P., & Benson, P. (2010). Categorical Perception of Facial Expressions: Categories and their Internal Structure. Cognition & Emotion, 11, 1–23. 10.1080/026999397380005

Gfeller, K., Olszewski, C., Turner, C., Gantz, B., & Oleson, J. (2006). Music Perception with Cochlear Implants and Residual Hearing. Audiology and Neurotology, 11(Suppl. 1), 12–15. 10.1159/000095608

Giannantonio, S., Polonenko, M. J., Papsin, B. C., Paludetti, G., & Gordon, K. A. (2015). Experience changes how emotion in music is judged: Evidence from children listening with bilateral cochlear implants, bimodal devices, and normal hearing. PLoS ONE, 10(8), 1–29. 10.1371/journal.pone.0136685

Glasberg, B. R., & Moore, B. C. J. (2002). A Model of Loudness Applicable to Time-Varying Sounds. Journal of the Audio Engineering Society, 50(5), 331–342.

Gomez, P., & Danuser, B. (2007). Relationships between musical structure and psychophysiological measures of emotion. - PsycNET. Emotion, 7(2), 377–387. 10.1037/1528-3542.7.2.377

Green, D. M., & Swets, J. A. (1988). Signal detection theory and psychophysics. (Reprint edition.). Peninsula.

Grewe, O., Nagel, F., Kopiez, R., & Altenmüller, E. (2005). How Does Music Arouse “Chills”? Annals of the New York Academy of Sciences, 2060(1), 446–449.

Hake, R., Bürgel, M., Nguyen, N. K., Greasley, A., Müllensiefen, D., & Siedenburg, K. (2023). Development of an adaptive test of musical scene analysis abilities for normal-hearing and hearing-impaired listeners. Behavior Research Methods. 10.3758/s13428-023-02279-y

Harding, E. E., Gaudrain, E., Hrycyk, I. J., Harris, R. L., Tillmann, B., Maat, B., Free, R. H., & Başkent, D. (2023). Musical Emotion Categorization with Vocoders of Varying Temporal and Spectral Content. Trends in Hearing, 27, 1–19. 10.1177/23312165221141142

Harrison, N. A., Wilson, C. E., & Critchley, H. D. (2007). Processing of observed pupil size modulates perception of sadness and predicts empathy. Emotion, 7(4), 724–729. 10.1037/1528-3542.7.4.724

Hess, U. (2017). Chapter 5—Emotion Categorization. In H. Cohen & C. Lefebvre (Red.), Handbook of Categorization in Cognitive Science (Second Edition) (pp. 107-126). Elsevier. 10.1016/B978-0-08-101107-2.00005-1

Hidalgo, C., Zécri, A., Pesnot-Lerousseau, J., Truy, E., Roman, S., Falk, S., Dalla Bella, S., & Schön, D. (2021). Rhythmic Abilities of Children With Hearing Loss. Ear and Hearing, 42(2), 364–372. 10.1097/AUD.0000000000000926

Hinderink, J. B., Krabbe, P. F. M., & Van Den Broek, P. (2000). Development and application of a health-related quality-of-life instrument for adults with cochlear implants: The Nijmegen Cochlear Implant Questionnaire. Otolaryngology - Head and Neck Surgery, 123(6), 756–765. 10.1067/mhn.2000.108203

Hoemann, K., Wu, R., LoBue, V., Oakes, L. M., Xu, F., & Barrett, L. F. (2020). Developing an Understanding of Emotion Categories: Lessons from Objects. Trends in Cognitive Sciences, 24(1), 39–51. 10.1016/j.tics.2019.10.010

Holden, L. K., Finley, C. C., Firszt, J. B., Holden, T. A., Brenner, C., Potts, L. G., Gotter, B. D., Vanderhoof, S. S., Mispagel, K., Heydebrand, G., & Skinner, M. W. (2013). Factors Affecting Open-Set Word Recognition in Adults With Cochlear Implants. Ear and Hearing, 34(3), 342. 10.1097/AUD.0b013e3182741aa7

Hopyan, T., Manno, F. A. M., Papsin, B. C., & Gordon, K. A. (2016). Sad and happy emotion discrimination in music by children with cochlear implants. Child Neuropsychology, 22(3), 366–380. 10.1080/09297049.2014.992400

Ilie, G., & Thompson, W. F. (2006). A Comparison of Acoustic Cues in Music and Speech for Three Dimensions of Affect. Music Perception, 23(4), 319–330. 10.1525/mp.2006.23.4.319

Jiam, N. T., Caldwell, M., Deroche, M. L., Chatterjee, M., & Limb, C. J. (2017). Voice emotion perception and production in cochlear implant users. Hearing Research, 352, 30–39. 10.1016/j.heares.2017.01.006

Knobloch, M., Verhey, J. L., Ziese, M., Nitschmann, M., Arens, C., & Böckmann-Barthel, M. (2018). Musical Harmony in Electric Hearing. Music Perception, 36(1), 40–52. 10.1525/mp.2018.36.1.40

Lassaletta, L., Castro, A., Bastarrica, M., Pérez-Mora, R., Herrán, B., Sanz, L., Josefa de Sarriá, M., & Gavilán, J. (2008). Musical Perception and Enjoyment in Post-Lingual Patients With Cochlear Implants. Acta Otorrinolaringologica (English Edition), 59(5), 228–234. 10.1016/s2173-5735(08)70228-x

Lawrence, M. A. (2016). Package ‘ez’. (Versie 4) [Software].

Lévêque, Y., Teyssier, P., Bouchet, P., Bigand, E., Caclin, A., & Tillmann, B. (2018). Musical emotions in congenital amusia: Impaired recognition, but preserved emotional intensity. Neuropsychology, 32(7), 880–894. 10.1037/neu0000461

Liégeois-Chauvel, C., Bénar, C., Krieg, J., Delbé, C., Chauvel, P., Giusiano, B., & Bigand, E. (2014). How functional coupling between the auditory cortex and the amygdala induces musical emotion: A single case study. Cortex, 60, 82–93. 10.1016/j.cortex.2014.06.002

Limb, C. J. (2006). Cochlear implant-mediated perception of music. Current Opinion in Otolaryngology & Head and Neck Surgery, 14(5), 337–340. 10.1097/01.moo.0000244192.59184.bd

Lin, Y.-S., Wu, C.-M., Limb, C. J., Lu, H.-P., Feng, I. J., Peng, S.-C., Deroche, M. L. D., & Chatterjee, M. (2022). Voice emotion recognition by Mandarin-speaking pediatric cochlear implant users in Taiwan. Laryngoscope Investigative Otolaryngology, 7(1), 250–258. 10.1002/lio2.732

Luo, X., Fu, Q.-J., & Galvin, J. J. (2007). Cochlear Implants Special Issue Article: Vocal Emotion Recognition by Normal-Hearing Listeners and Cochlear Implant Users. Trends in Amplification, 11(4), 301–315. 10.1177/1084713807305301

Luo, X., Kern, A., & Pulling, K. R. (2018). Vocal emotion recognition performance predicts the quality of life in adult cochlear implant users. The Journal of the Acoustical Society of America, 144(5), EL429-EL435. 10.1121/1.5079575

Macherey, O., & Carlyon, R. P. (2014). Cochlear implants. Current Biology, 24(18), R878–R884. 10.1016/j.cub.2014.06.053

Macmillan, N. A., & Creelman, C. D. (2004). Detection Theory: A User’s Guide. Psychology Press.

McDermott, H. J. (2004). Music Perception with Cochlear Implants: A Review. Trends in Amplification, 8(2), 49-82. 10.1177/108471380400800203

Morise, M., Yokomori, F., & Ozawa, K. (2016). WORLD: A Vocoder-Based High-Quality Speech Synthesis System for Real-Time Applications. IEICE Transactions on Information and Systems, E99.D(7), 1877-1884. 10.1587/transinf.2015EDP7457

Müller, M. (2021). Chord Recognition. In M. Müller (Red.), Fundamentals of Music Processing: Using Python and Jupyter Notebooks (pp. 241-308). Springer International Publishing. 10.1007/978-3-030-69808-9_5

Nagels, L., Gaudrain, E., Vickers, D., Lopes, M. M., Hendriks, P., & Başkent, D. (2020). Development of vocal emotion recognition in school-age children: The EmoHI test for hearing-impaired populations. PeerJ, 8, e8773. 10.7717/peerj.8773

Nieminen, S., Istók, E., Brattico, E., & Tervaniemi, M. (2012). The development of the aesthetic experience of music: Preference, emotions, and beauty. Musicae Scientiae, 16(3), 372–391. 10.1177/1029864912450454

Nussbaum, C., Schirmer, A., & Schweinberger, S. R. (2022). Contributions of fundamental frequency and timbre to vocal emotion perception and their electrophysiological correlates. Social Cognitive and Affective Neuroscience, 17(12), 1145–1154. 10.1093/scan/nsac033

Nyirjesy, S. C., Lewis, J. H., Hallak, D., Conroy, S., Moberly, A. C., & Tamati, T. N. (2023). Evaluating Listening Effort in Unilateral, Bimodal, and Bilateral Cochlear Implant Users. Otolaryngology--Head and Neck Surgery: Official Journal of American Academy of Otolaryngology-Head and Neck Surgery. 10.1002/ohn.609

Paquette, S., Ahmed, G. D., Goffi-Gomez, M. V., Hoshino, A. C. H., Peretz, I., & Lehmann, A. (2018). Musical and vocal emotion perception for cochlear implants users. Hearing Research, 370, 272–282. 10.1016/j.heares.2018.08.009

Park, M., Gutyrchik, E., Welker, L., Carl, P., Pöppel, E., Zaytseva, Y., Meindl, T., Blautzik, J., Reiser, M., & Bao, Y. (2015). Sadness is unique: Neural processing of emotions in speech prosody in musicians and non-musicians. Frontiers in Human Neuroscience, 8. 10.3389/fnhum.2014.01049

Patterson, R. D., Walters, T. C., Monaghan, J. J. M., & Gaudrain, E. (2010). Reviewing the Definition of Timbre as it Pertains to the Perception of Speech and Musical Sounds. In E. A. Lopez-Poveda, A. R. Palmer, & R. Meddis (Red.), The Neurophysiological Bases of Auditory Perception (pp. 223-233). Springer. 10.1007/978-1-4419-5686-6_21

Phillips, M. L., Drevets, W. C., Rauch, S. L., & Lane, R. (2003). Neurobiology of emotion perception I: The neural basis of normal emotion perception. Biological Psychiatry, 54(5), 504–514. 10.1016/s0006-3223(03)00168-9

Plant, K., McDermott, H., van Hoesel, R., Dawson, P., & Cowan, R. (2016). Factors Predicting Postoperative Unilateral and Bilateral Speech Recognition in Adult Cochlear Implant Recipients with Acoustic Hearing. Ear and Hearing, 37(2), 153–163. 10.1097/AUD.0000000000000233

Pralus, A., Belfi, A., Hirel, C., Lévêque, Y., Fornoni, L., Bigand, E., Jung, J., Tranel, D., Nighoghossian, N., Tillmann, B., & Caclin, A. (2020). Recognition of musical emotions and their perceived intensity after unilateral brain damage. Cortex, 130, 78–93. 10.1016/j.cortex.2020.05.015

Rachman, L., Babaoğlu, G., Özkişi Yazgan, B., Ertürk, P., Gaudrain, E., Nagels, L., Launer, S., Derleth, P., Singh, G., Jehle, F., Chatterjee, M., Yücel, E., Sennaroğlu, G., & Başkent, D. (submitted).

Russell, J. A. (1980). A circumplex model of affect. Journal of Personality and Social Psychology, 39(6), 1161–1178.

Saliba, J., Bortfeld, H., Levitin, D. J., & Oghalai, J. S. (2016). Functional near-infrared spectroscopy for neuroimaging in cochlear implant recipients. Hearing research, 338, 64–75. 10.1016/j.heares.2016.02.005

Schlosberg, H. (1952). The Description of Facial Expressions in Terms of Two Dimensions. Journal of Experimental Psychology, 44(4), 229–237.

Schorr, E. A., Roth, F. P., & Fox, N. A. (2009). Quality of Life for Children With Cochlear Implants: Perceived Benefits and Problems and the Perception of Single Words and Emotional Sounds. Journal of Speech, Language, and Hearing Research, 52(1), 141–152. 10.1044/1092-4388(2008/07-0213)

Schubert, E. (2004). Modeling Perceived Emotion With Continuous Musical Features. Music Perception, 21(4), 561–585. 10.1525/mp.2004.21.4.561

Shannon, R. V. (1983). Multichannel electrical stimulation of the auditory nerve in man. II. Channel interaction. Hearing Research, 12(1), 1–16. 10.1016/0378-5955(83)90115-6

Snel-Bongers, J., Netten, A. P., Boermans, P.-P. B. M., Rotteveel, L. J. C., Briaire, J. J., & Frijns, J. H. M. (2018). Evidence-Based Inclusion Criteria for Cochlear Implantation in Patients With Postlingual Deafness. Ear and Hearing, 39(5), 1008. 10.1097/AUD.0000000000000568

Souza, P., Arehart, K., Miller, C. W., & Muralimanohar, R. K. (2011). Effects of Age on F0 Discrimination and Intonation Perception in Simulated Electric and Electroacoustic Hearing. Ear and Hearing, 32(1), 75. 10.1097/AUD.0b013e3181eccfe9

van Dijk, J. E., van Olphen, A. F., Langereis, M. C., Mens, L. H., Brokx, J. P., & Smoorenburg, G. F. (1999). Predictors of cochlear implant performance. Audiology: Official Organ of the International Society of Audiology, 38(2), 109–116. 10.3109/00206099909073010

Velde, D. J. V. D., Schiller, N. O., Levelt, C. C., Heuven, V. J. V., Beers, M., Briaire, J. J., & Frijns, J. H. M. (2019). Prosody perception and production by children with cochlear implants. Journal of Child Language, 46(1), 111–141. 10.1017/S0305000918000387

Vieillard, S., Peretz, I., Gosselin, N., Khalfa, S., Gagnon, L., & Bouchard, B. (2008). Happy, sad, scary and peaceful musical excerpts for research on emotions. Cognition & Emotion, 22(4), 720–752. 10.1080/02699930701503567

Volkova, A., Trehub, S. E., Glenn Schellenberg, E., Papsin, B. C., & Gordon, K. A. (2013). Children with bilateral cochlear implants identify emotion in speech and music. Cochlear Implants International, 14(2), 80–91. 10.1179/1754762812Y.0000000004

Von Eiff, C. I., Frühholz, S., Korth, D., Guntinas-Lichius, O., & Schweinberger, S. R. (2022). Crossmodal benefits to vocal emotion perception in cochlear implant users. iScience, 25(12), 105711. 10.1016/j.isci.2022.105711

von Eiff, C. I., Skuk, V. G., Zäske, R., Nussbaum, C., Frühholz, S., Feuer, U., Guntinas-Lichius, O., & Schweinberger, S. R. (2022). Parameter-Specific Morphing Reveals Contributions of Timbre to the Perception of Vocal Emotions in Cochlear Implant Users. Ear and Hearing, 43(4), 1178. 10.1097/AUD.0000000000001181

Wang, X., Wei, Y., & Yang, D. (2022). Cross-cultural analysis of the correlation between musical elements and emotion. Cognitive Computation and Systems, 4(2), 116–129. 10.1049/ccs2.12032

Widen, S. C., & Russell, J. A. (2004). The relative power of an emotion’s facial expression, label, and behavioral consequence to evoke preschoolers’ knowledge of its cause. Cognitive Development, 19(1), 111–125. 10.1016/j.cogdev.2003.11.004

Zaltz, Y., & Kishon-Rabin, L. (2022). Difficulties Experienced by Older Listeners in Utilizing Voice Cues for Speaker Discrimination. Frontiers in Psychology, 13. https://www.frontiersin.org/articles/10.3389/fpsyg.2022.797422

Zimmer, V., Verhey, J. L., Ziese, M., & Böckmann-Barthel, M. (2019). Harmony Perception in Prelingually Deaf, Juvenile Cochlear Implant Users. Frontiers in Neuroscience, 13, 466. 10.3389/fnins.2019.00466

