## Supplementary material for "Vocal and musical emotion perception, voice cue discrimination, and quality of life in cochlear implant users with and without acoustic hearing": Confidence ratings are reported in supplementary materials S1.

#### S1 Confidence ratings

##### Analyses

Confidence ratings were collected after each trial, and scaled to the mean and sd of each participant per emotion category before being correlated with d' (Pearsons, 2-tailed). Confidence ratings from both correct and incorrect categorization trials were used. Confidence ratings were entered into the same ANOVA as sensitivity to assess any differences between confidence and d'. To see whether sensitivity increased as confidence to make the categorization increased, confidence ratings were further entered into correlations with sensitivity for each presented emotion category.

##### Results

Confidence rating results were as follows: joy, mean = 58.47, *SD* = 15.03; fear, mean = 53.48, *SD* = 15.36; serenity, mean = 58.98, *SD* = 14.62; sadness, mean = 54.03, *SD* = 14.83. Confidence ratings in all categories were significantly above "Very little confidence" ($t$'s > 20.6, $p$'s < .0001). A one-way ANOVA yielded a main effect of Presented Emotion Category ($F(3,81)=7.78$, $p=.0001$, $\eta^2_G=.036$). However, posthoc pairwise t-tests did not show any significant differences between any categories. When d' and confidence ratings were correlated across participants for each emotion category, no correlations were significant for any emotion (all $p_{adj}$'s > .615, $N$ = 28), indicating that confidence had no statistical link with sensitivity.

An ANOVA with added factor Acoustic hearing was conducted with no main effect of Acoustic hearing ($F$(2,25)=0.004, $p$=1.0, $\eta^2_G$=0.0003), indicating that confidence ratings did not differ between unilateral and bimodal participants. No correlations between confidence ratings and d' per emotion category were significant within either group (all $p_{adj}$'s > .18).

#### Discussion

Sensitivity diverged from confidence ratings; interestingly the positive emotion categories joy and peace were more confidently assessed while sensitivity was highest in categories joy and fear. Considering the mean age of our study falls in an 'older adult' category, this result could reflect an older adult 'positivity bias' found previously when categorizing emotions from musical excerpts (Lima & Castro, 2011; Pearce & Halpern, 2015). In both of those studies, the rating of emotions in the presented category resulted in older adults generally rating sad and fearful categories as less sad and fearful compared to younger adults, while happy and peaceful categories were more similarly rated across age groups. Thus in our study, the confidence of positive-valence categories may still be a general reflection of older adults' tendency to 'look on the bright side' as a general emotion regulation skill (Mather & Carstensen, 2005).

### References

Lima, C. F., & Castro, S. L. (2011). Speaking to the trained ear: musical expertise enhances the recognition of emotions in speech prosody. *Emotion*, *11*(5), 1021.

Mather, M., & Carstensen, L. L. (2005). Aging and motivated cognition: The positivity effect in attention and memory. *Trends in cognitive sciences*, *9*(10), 496-502.

Pearce, M. T., & Halpern, A. R. (2015). Age-related patterns in emotions evoked by music. *Psychology of Aesthetics, Creativity, and the Arts*, *9*(3), 248.
